# Moving towards precision post-stroke rehabilitation: A systematic review and meta-analysis of wearable sensor-derived walking activities in daily living

**DOI:** 10.1101/2025.10.23.25338670

**Authors:** Joy C. Ezeugwa, Aiza Khan, Deborah Okunsanya, Liz Dennett, Brian H. Buck, Patricia J. Manns, Victor E. Ezeugwu

## Abstract

**Background:** Physical rehabilitation interventions can enhance functional capacity after stroke, but improved ability doesn’t always lead to better real-world performance. Understanding this gap is key to optimizing post-stroke recovery strategies.

**Objectives:** This research aimed to evaluate the effectiveness of physical rehabilitation interventions specifically exercise, behavior change techniques (BCTs), or their combination on real-world walking measured using wearable sensors and capacity (gait speed and walking endurance) outcomes in stroke survivors.

**Methods:** This systematic review and meta-analysis followed PRISMA guidelines. Comprehensive searches were conducted in Medline, Embase, CINAHL, and Scopus up to January 2025. Randomized controlled trials involving stroke survivors receiving physical rehabilitation interventions—exercise, BCTs, or both—compared to exercise-only or usual care were included. Outcomes assessed were daily steps, gait speed (comfortable and fastest), and endurance (6-minute walk test). Meta-analyses using random-effects models (STATA 18) reported standardized mean differences (SMDs) and 95% confidence intervals (CIs). Heterogeneity was evaluated using I² statistics.

**Results:** Of 1,782 references screened, 28 studies met the inclusion criteria, and 23 were included in meta-analyses comprising 2,327 participants. Exercise-only interventions produced a small but significant improvement in daily steps (SMD = 0.23; 95% CI: 0.03 to 0.44; I^2^ = 36.6%; *moderate certainty*), and moderate improvements in both comfortable gait speed (SMD = 0.38; 95% CI: 0.19 to 0.57; I^2^ = 35.5%; *moderate certainty*) and endurance (SMD = 0.39; 95% CI: 0.26 to 0.52; I^2^ = 0%; *moderate certainty*). BCT-only interventions demonstrated a larger effect on daily steps (SMD = 0.41; 95% CI: 0.19 to 0.63; I^2^ = 0%; moderate certainty). In contrast, combined exercise and BCT interventions did not yield significant improvements in any outcomes and were supported by *very low to low certainty* of evidence.

**Conclusion:** Exercise-only interventions improve gait speed and endurance after stroke, with small gains in daily steps. BCT-only interventions yield greater improvements in daily walking activity. Combined interventions show limited added benefit.

**Protocol registration:** This study has been registered in PROSPERO (No. CRD42023411679)

## 1. Introduction

Stroke remains a leading global health issue, affecting over 25 million individuals worldwide as of 2013 (1). A significant proportion of stroke survivors experience physical and psychological impairments that may hinder their ability to engage in physical activity (PA) (2,3). Stroke recurrence is common, with nearly one in four stroke survivors likely to have a recurrence (4), underscoring the importance of mitigating modifiable risk factors such as physical inactivity and prolonged sedentary behavior (SB; sitting/lying activities requiring ≤1.5 metabolic equivalent tasks (METs)) (5). Physical inactivity and prolonged SB are associated with higher cardiovascular risk and may contribute to stroke recurrence and mortality (6–13).

Current physical activity and exercise guidelines for stroke survivors include 20 to 60 minutes of aerobic exercise three to five days per week, in addition to strength, and flexibility training two to three days weekly (2,14). These guidelines are designed to enhance stroke recovery and to prevent secondary stroke events. Despite these well-defined recommendations, evidence consistently shows that stroke survivors are physically inactive and engage in prolonged SB (3,15,16). Walking is a simple, accessible, and commonly recommended form of PA after stroke. Daily walking, commonly measured as steps per day, has been linked to numerous benefits including lowered blood pressure, weight reduction, and decreased cardiovascular disease risk (17–19). Walking activity is typically measured in two ways: *capacity (e.g., walking speed, endurance)*—the maximum walking ability measured under standardized conditions—and *performance (e.g., daily steps)*—how much walking occurs in daily life (20). Most rehabilitation programs emphasize improving walking capacity, such as through speed and distance training in clinical settings (21–23). However, growing evidence suggests that gains in capacity do not necessarily translate into improved real-world performance, such as increased daily steps (3,24,25). This disconnect highlights the need to investigate which interventions may improve capacity and performance outcomes after stroke.

Behavioral interventions have emerged as promising strategies to bridge the gap between physical capacity and real-world activity after stroke (17). Techniques such as daily step monitoring, feedback on behaviour, and personalized coaching have shown success in promoting sustained PA, especially once patients transition back into community settings after discharge (26,27). Notably, exercise-only interventions often fall short, while those incorporating behavior change techniques (BCTs) lead to more substantial gains in daily step counts (28). For example, one RCT showed that participants using step activity monitoring (SAM) increased their steps by over 1,500 per day, compared to 406 steps/day in the exercise-only group (17). This finding suggests that interventions using BCTs may improve performance outcomes after stroke.

While previous systematic reviews have examined PA interventions post-stroke (28–35), none has directly compared the effectiveness of rehabilitation interventions on both performance and capacity outcomes. These reviews have also been limited by heterogeneity and wide variability in study designs.

The current systematic review and meta-analysis aim to synthesize evidence on physical rehabilitation interventions, including exercise-only, BCT-only, and combined approaches, on performance and capacity outcomes post-stroke. Our results may strengthen the evidence base on which physical rehabilitation interventions most effectively improve these outcomes, thereby informing the development and implementation of targeted rehabilitation strategies.

## 2. Methods

### Design

Systematic Review and Meta-Analysis of Randomized Controlled Trials This systematic review adhered to the Preferred Reporting Items for Systematic reviews and Meta-Analyses (PRISMA) guideline (http://www.prisma-statement.org) (S1 Table PRISMA Checklist). The study protocol was prospectively registered on PROSPERO (International Prospective Register of Ongoing Systematic Reviews - CRD42023411679) (https://www.crd.york.ac.uk/PROSPERO/view/CRD42023411679).

### Deviations from protocol

There were some deviations from our registered protocol on PROSPERO. Initially titled *“Moving towards precision post-stroke rehabilitation guided by wearable sensors,”* the protocol was slightly modified to align with the current review title. The originally registered outcomes included daily steps, endurance, sedentary time, sleep time, gait speed (10-meter walk test), mobility (Timed Up and Go test - TUG), and global disability (modified Rankin Scale - mRS). However, TUG and mRS were excluded from the final review as they do not directly measure physical performance or capacity, but rather assess functional mobility and global disability, respectively.

### Search Strategy

A comprehensive literature search was conducted across Medline, Embase, CINAHL, and Scopus for English-language articles published up to March 7, 2023, with an updated search completed in January 2025. The review aimed to evaluate the impact of physical rehabilitation approaches on physical function outcomes specifically capacity and performance following stroke. The search and selection process were guided by the PICO framework: the Population included stroke survivors; Interventions comprised exercise-only, behaviour change techniques (BCT)-only, or combined approaches; the Comparator was standard care or low-intensity exercise; and the Outcomes focused on physical performance (measured by daily step count) and capacity (measured by the 6-minute walk test and gait speed). Following the literature search, identified articles were imported into Covidence (Veritas Health Innovation, Melbourne, Australia), where duplicates were removed. Two independent reviewers screened all titles and abstracts, and full-text articles for eligibility. Any conflicts arising during the screening phases were resolved by a third independent reviewer.

The following search strings were utilized: (Stroke or poststroke or cerebrovascular disease or cerebrovascular accident* or ischemic attack*) AND (FitBit* or Smartwatch* “Apple Watch*” or galaxy-watch* or Polar-H*) AND (Actigraph* or ActivPAL or ambulatory monitor* or gyroscope* or pedometer* or pedometer* or gps or global positioning system* or fitness-tracker* or activity-tracker* or accelerometer* or accelerometre*) AND (Movement behavior* or sedentary or sleep or physical activity or exercis* or gait or walk or walking or step or steps AND randomized controlled trials. A detailed account of the search strategy, including exact search strings, is provided as **S1** File. The Evidence-Based Review of Stroke Rehabilitation (www.ebrsr.com) was cross-referenced to ensure comprehensive article identification. This resource was used to supplement the database search by identifying key studies and systematic reviews related to physical rehabilitation and functional outcomes post-stroke that may not have been captured through traditional database indexing.

### Study Selection

#### Inclusion Criteria

Studies were included in this systematic review if they met the following criteria: i) adults living with stroke aged ≥18 years, ii) any type of physical rehabilitation interventions, iii) the control group engaged in usual care or another type/different intensity of physical rehabilitation intervention, and iv) only randomized controlled trials (RCTs) were included. Outcomes included performance (steps/day) and capacity (comfortable/fastest gait speed and 6-minute walk distance) measures.

#### Exclusion Criteria

Studies were excluded if they did not meet the specified requirements. This included studies not written in English, systematic literature reviews, cross-sectional studies, cohort studies, or any other non-randomized clinical trials. Protocol studies without any data or results were also excluded. Additionally, studies were deemed ineligible if they used wearable technology exclusively as a treatment modality, solely focused on upper extremity function and mobility, or were conducted on individuals aged 18 years or younger. Finally, studies that solely used “exoskeletons” or “robotics” were excluded as these are not routine physical rehabilitation interventions.

### Data Extraction

Data extraction was conducted independently by two authors using a predetermined template created within the Covidence software. Data extracted included comprehensive information on various aspects of the studies: author(s) names, title and year of publication, number of subjects at the beginning and end of the intervention, demographic information (age, sex, time since stroke, type of stroke, severity of stroke), detailed intervention and control protocol (FITT-frequency, intensity, time, type), data collection environment (lab-based, community, etc.), type(s) of wearable device used, location of the device on the participant’s body, gait variable(s) or parameters examined, and primary outcomes (steps/day, endurance, and gait speed). Outcomes were reviewed at the endpoint or follow-up. For studies with multiple follow-up periods, the end of intervention data was extracted. Additionally, where applicable, statistical analyses such as p-values and correlational values were extracted.

### Quality Assessment: Risk of Bias

To assess the methodological quality of the included articles, a risk of bias assessment was performed by two independent reviewers using the Physiotherapy Evidence Database (PEDro) scale for RCTs (36). The PEDro scale is an 11-item checklist used to assess the quality of clinical trials, evaluating both the external and internal validity of the reviewed randomized clinical trial, as well as statistical reporting. Reviewers scored each item with a “yes” or “no” based on whether it was clearly stated in the article. A score of 9–10 indicates “excellent quality” with very high internal validity.

Scores between 6–8 are considered “good”, reflecting acceptable validity and only minor bias. A “fair” rating is assigned to studies scoring 4–5, suggesting moderate risk of bias and the need for cautious interpretation. Studies scoring below 4 are rated “poor”, indicating a high risk of bias and limited reliability of the findings (36).

### Quality Assessment: Certainty of Evidence

The certainty of evidence for each pooled effect was assessed using the Grading of Recommendations Assessment, Development and Evaluation (GRADE) approach (37). The GRADE system classifies the certainty of evidence into four distinct levels: *High*, meaning we are very confident that the true effect is close to the estimate; *Moderate*, meaning the true effect is likely close to the estimate, but may be substantially different; *Low*, meaning the true effect may be substantially different from the estimate; and *Very Low*, meaning the true effect is likely to be substantially different from the estimate (37).

Since all evidence originated from randomized controlled trials, the initial certainty for every outcome was rated as High. This certainty was systematically assessed and potentially downgraded across five domains: Risk of Bias, Inconsistency, Indirectness, Imprecision, and Publication Bias (37). Risk of Bias was evaluated using the PEDro scale, and a downgrade of one level was applied if methodological limitations were deemed a serious concern, such as the inclusion of multiple studies with PEDro scores of 6 or below. Inconsistency (heterogeneity) was measured using the I^2^ statistic and was downgraded one level if heterogeneity was substantial (I^2^ ≥ 50%) and the effect estimate was poorly defined.

Imprecision led to a one-level downgrade if the 95% Confidence Interval (CI) was wide and crossed the line of no effect, SMD =0, indicating uncertainty regarding the true clinical effect. No downgrades were applied for Indirectness or Publication Bias, as all evidence was judged to be direct and no bias was identified. Pooled effects are presented as the Standardized Mean Difference (SMD) with its 95% CI. For clinical interpretability, the SMD was converted to an estimated Mean Difference (MD) by multiplying the SMD by the control group’s standard deviation of the highest-weighted study. This estimated MD was then compared against the established Minimal Clinically Important Difference (MCID) for each outcome to assess clinical relevance.

### Meta-analysis

Meta-analysis was performed to analyze the between-group differences in the post-intervention outcomes that were reported in at least two studies. For each meta-analysis, data were pooled using random effects models to account for heterogeneity across studies, ensuring a more robust and conservative estimate for the pooled effect size. SMD and 95% CI were reported as the effect size for all meta-analyses to enable comparison of our outcomes: daily steps, comfortable gait speed, fastest gait speed, and walking endurance. Heterogeneity across studies was estimated using the I^2^ statistic (38) and classified as low (<30%), moderate (30%-50%), substantial (50%-75%), or considerable (>75%) (38). Studies that reported median and interquartile range were excluded from the meta-analysis but included in the descriptive review. When studies included more than one relevant intervention arm, group interventions were included as separate comparisons (e.g., 2024a, 2024b) within the meta-analysis. Findings are presented using forest plots and tables. Effect sizes (i.e., SMD) were classified following Cohen’s magnitude criteria for rehabilitation treatment effects (39): d=0.14 to 0.31 for a small effect size; d=0.31 to 0.61 for a medium effect size; and d>0.61 for a large effect size (39).

## 3. Results

### Study selection

A total of 1782 articles were identified (1764 from databases, 18 via citation searching). After removing duplicates, 877 were screened, and 79 full texts assessed. Of these, 51 were excluded due to study design (n=31), intervention (n=10), outcome (n=9), or duplication (n=1). Finally, a total of 28 studies met the inclusion criteria for qualitative synthesis (17,25,40–65), of which 23 studies were included in meta-analysis (17,25,40–43,45,47–58,60,62,63,65). The flow chart for study selection is shown in Figure 1.

**Figure 1.**
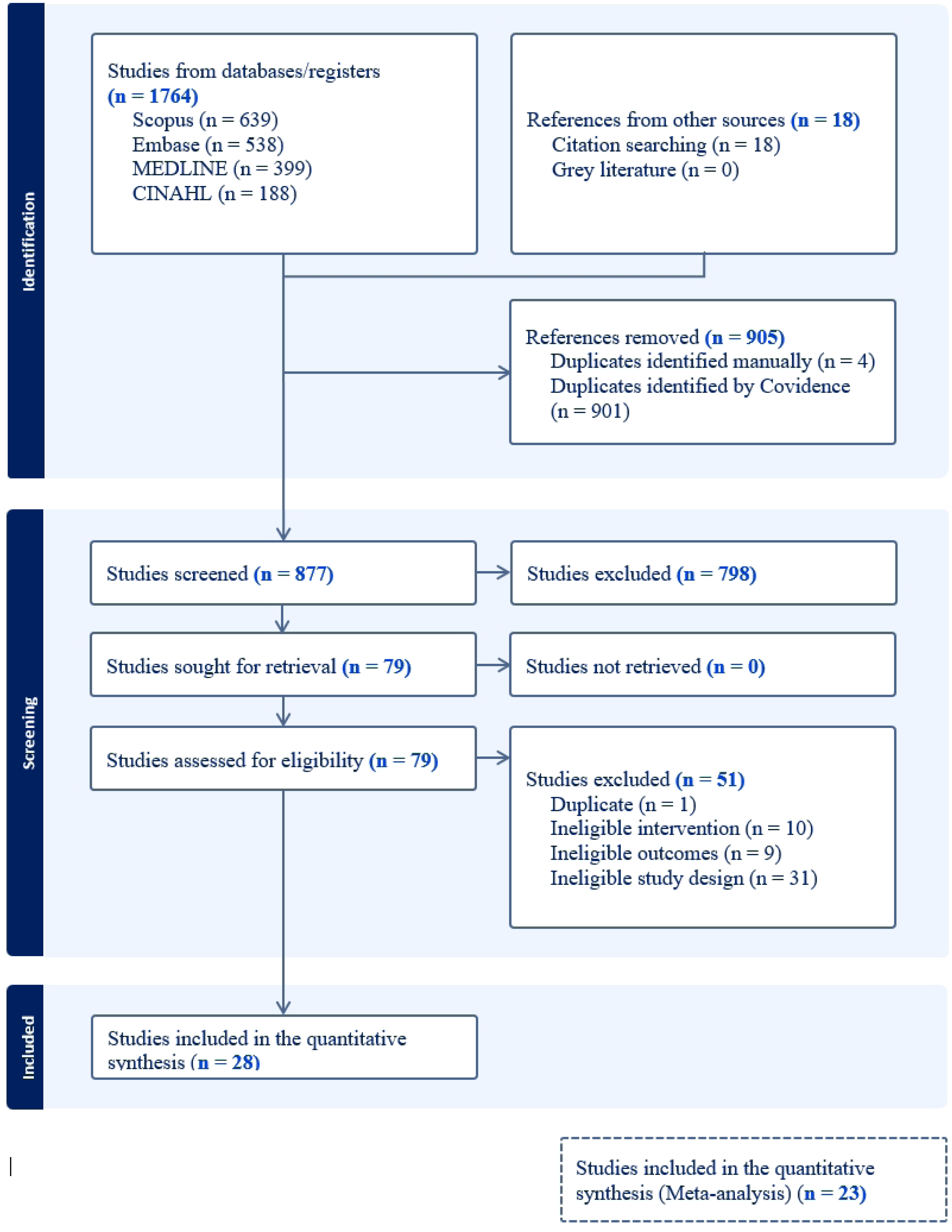
PRISMA flowchart of included trials

### Study Characteristics

The included studies were published between 2009 and 2024. Geographically, 8 studies were conducted in the United States (17,43–45,58,60,61,64), 4 in Japan (40,41,47,49), 2 in Australia (42,46), and one study each in Canada (48), Denmark (51), New Zealand (25), the Netherlands (53), Singapore (56), Belgium (59), Israel (62), and the United Kingdom (65). A total of 2,327 participants were included in the synthesis, with 1,311 allocated to intervention groups and 1,061 to control groups. Among them, 889 were female (512 in intervention groups and 377 in control groups). A detailed summary of the study characteristics is presented in Table 1.

**Table 1:**
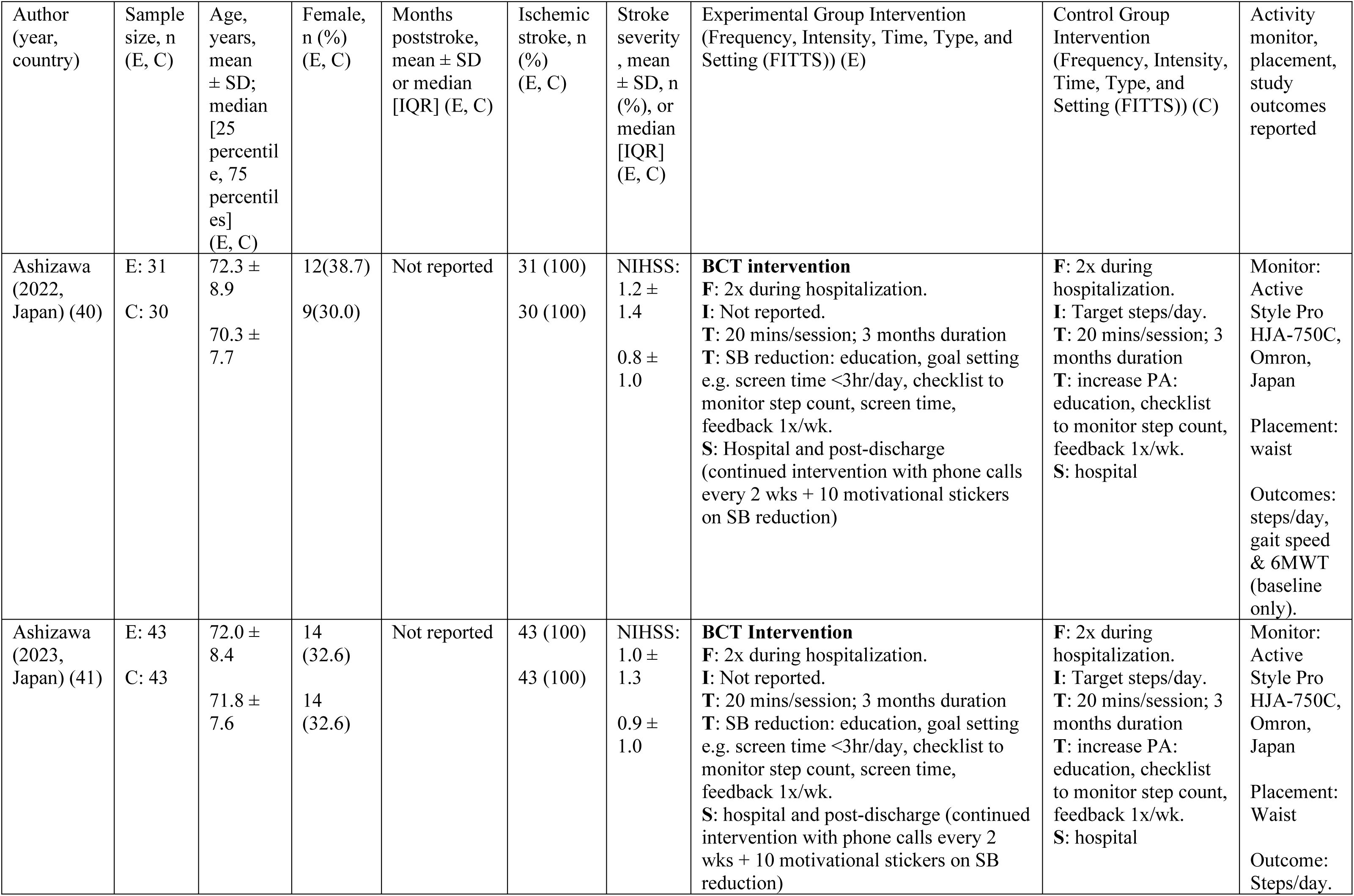
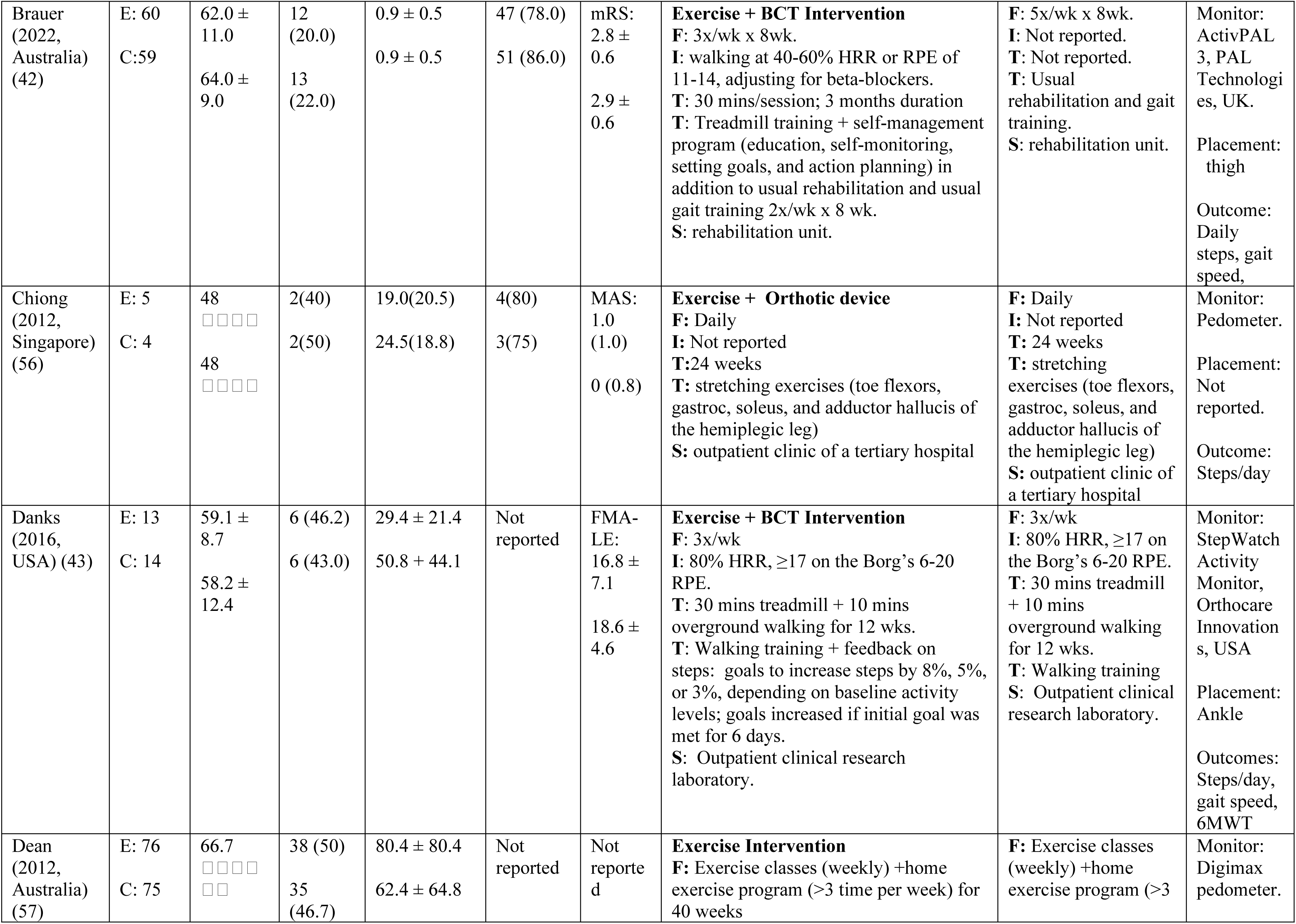
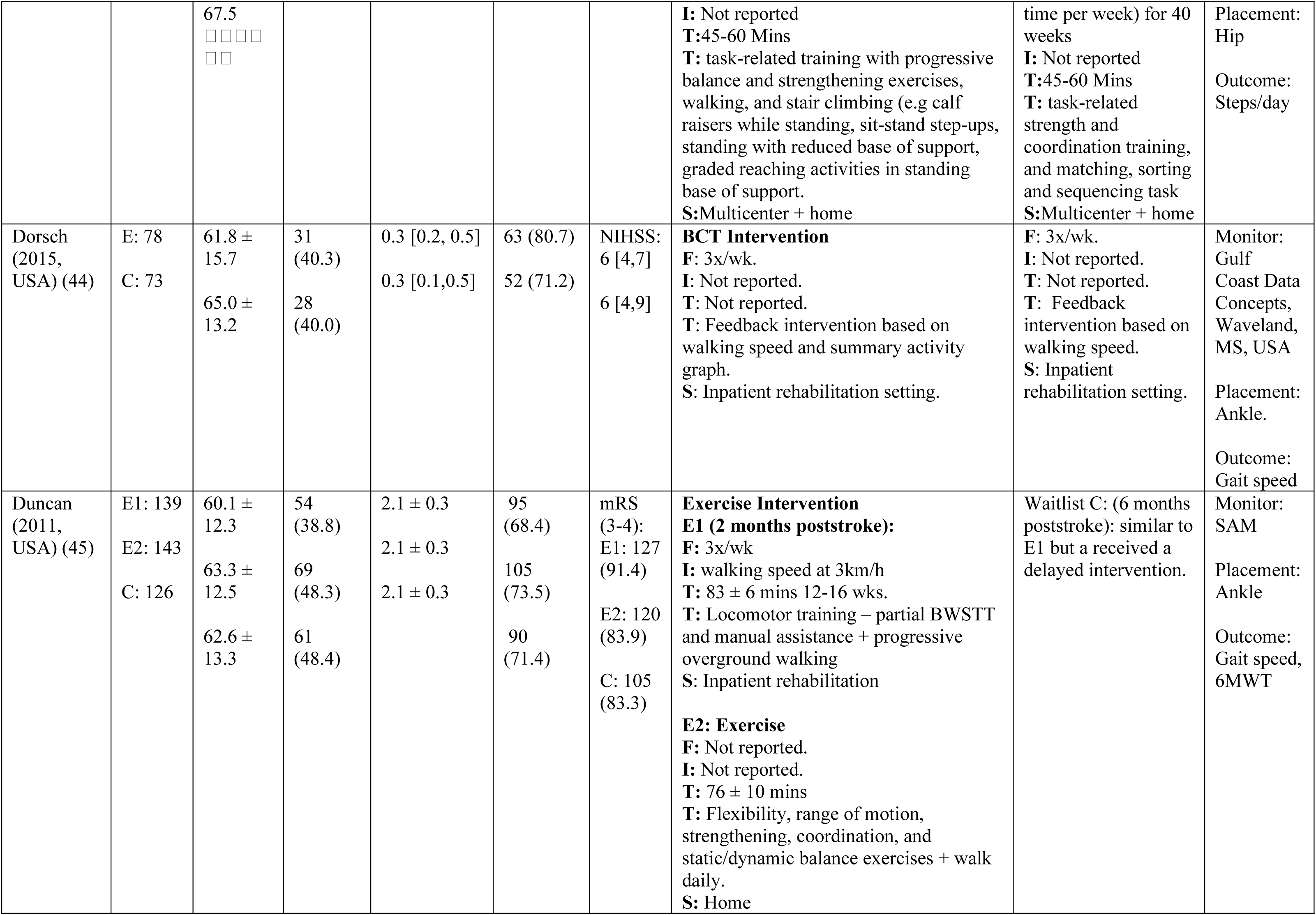
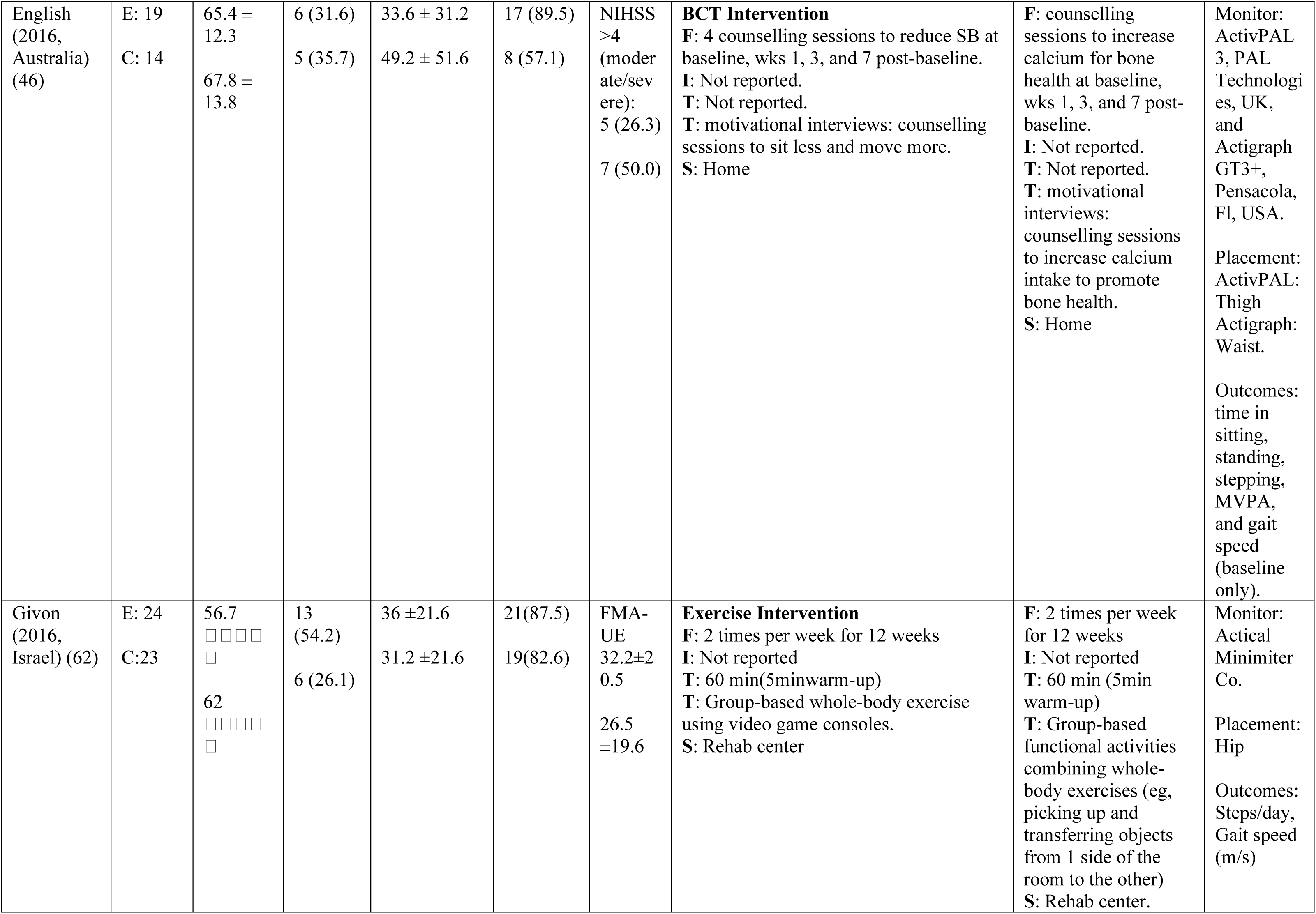
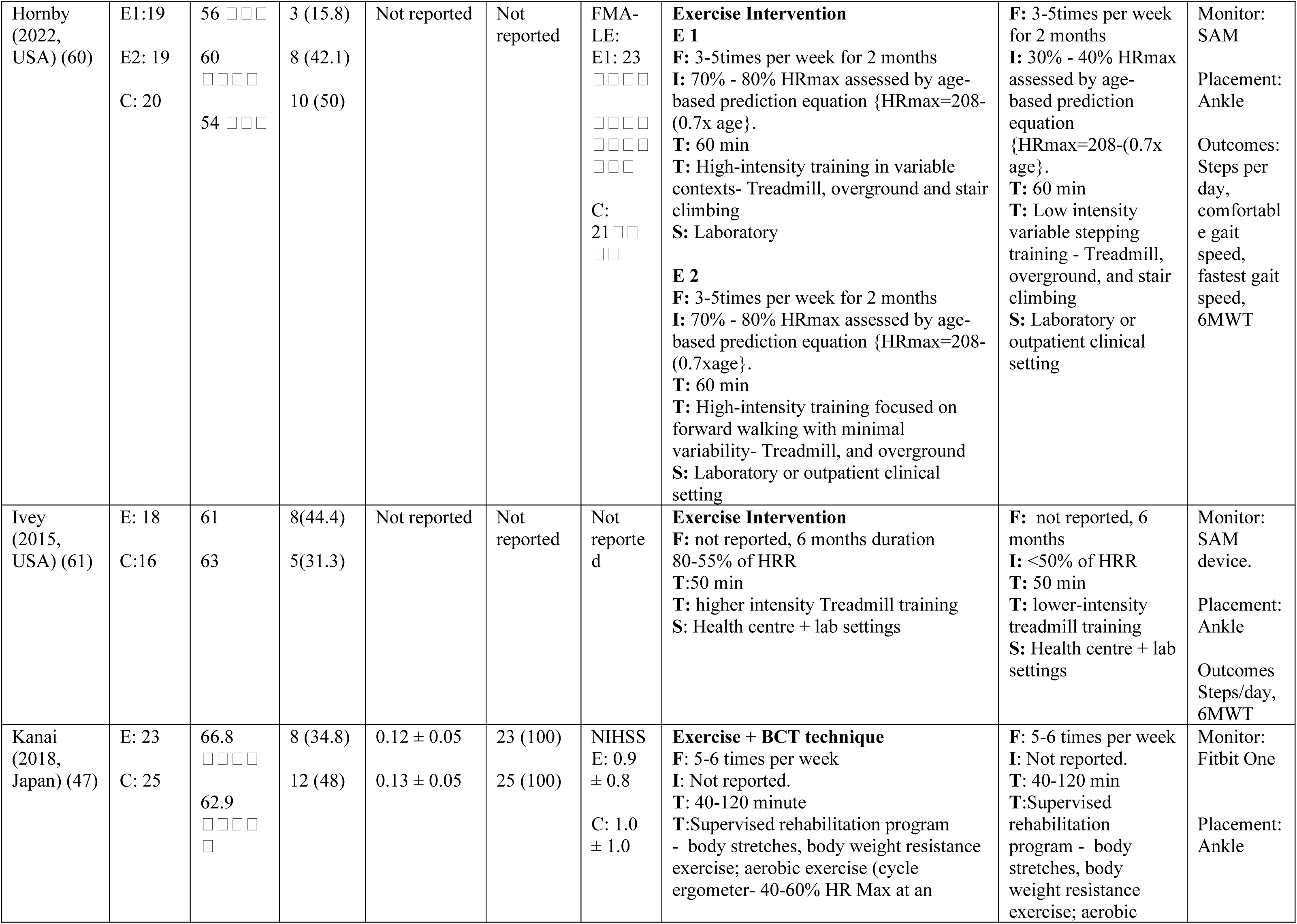
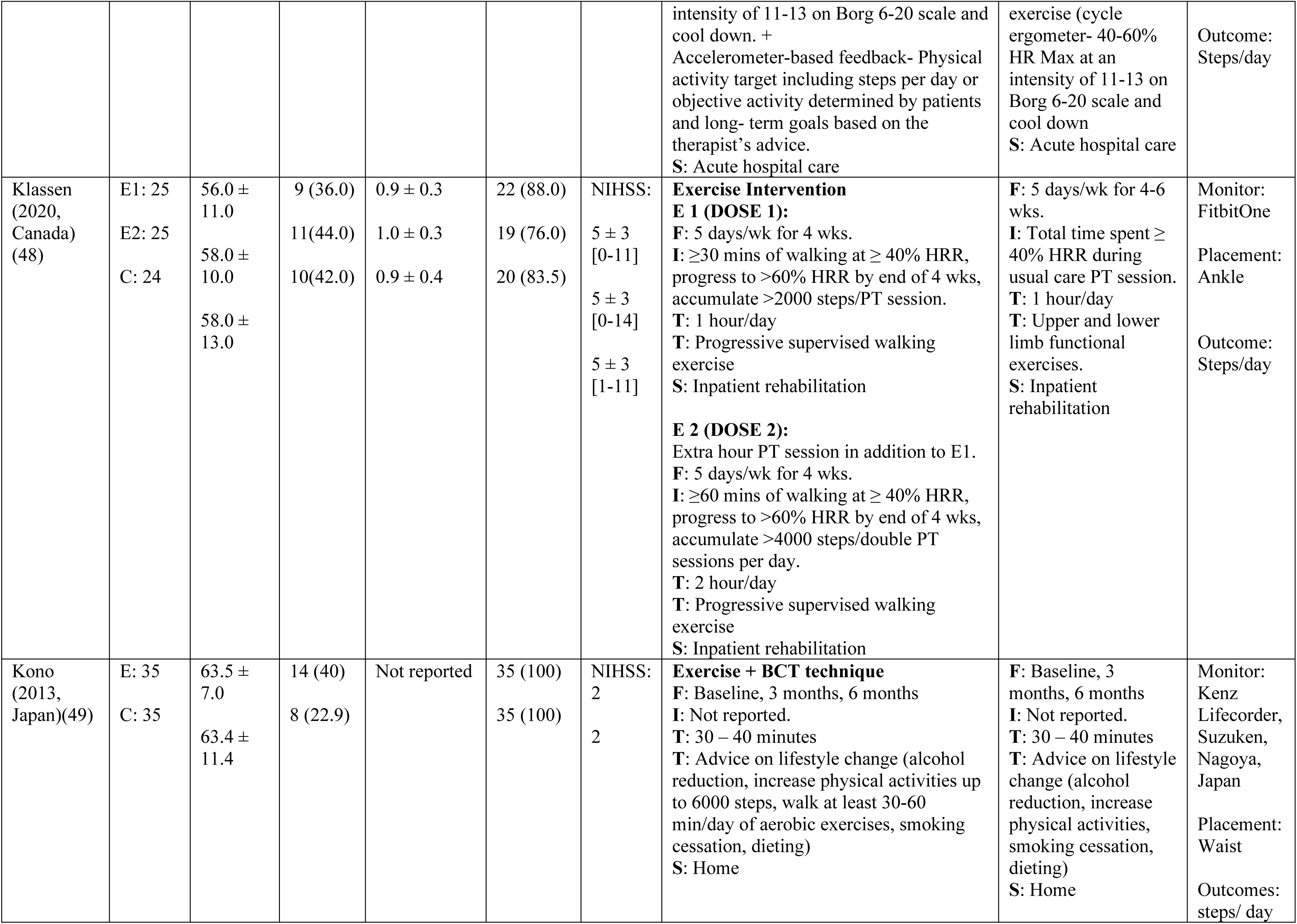
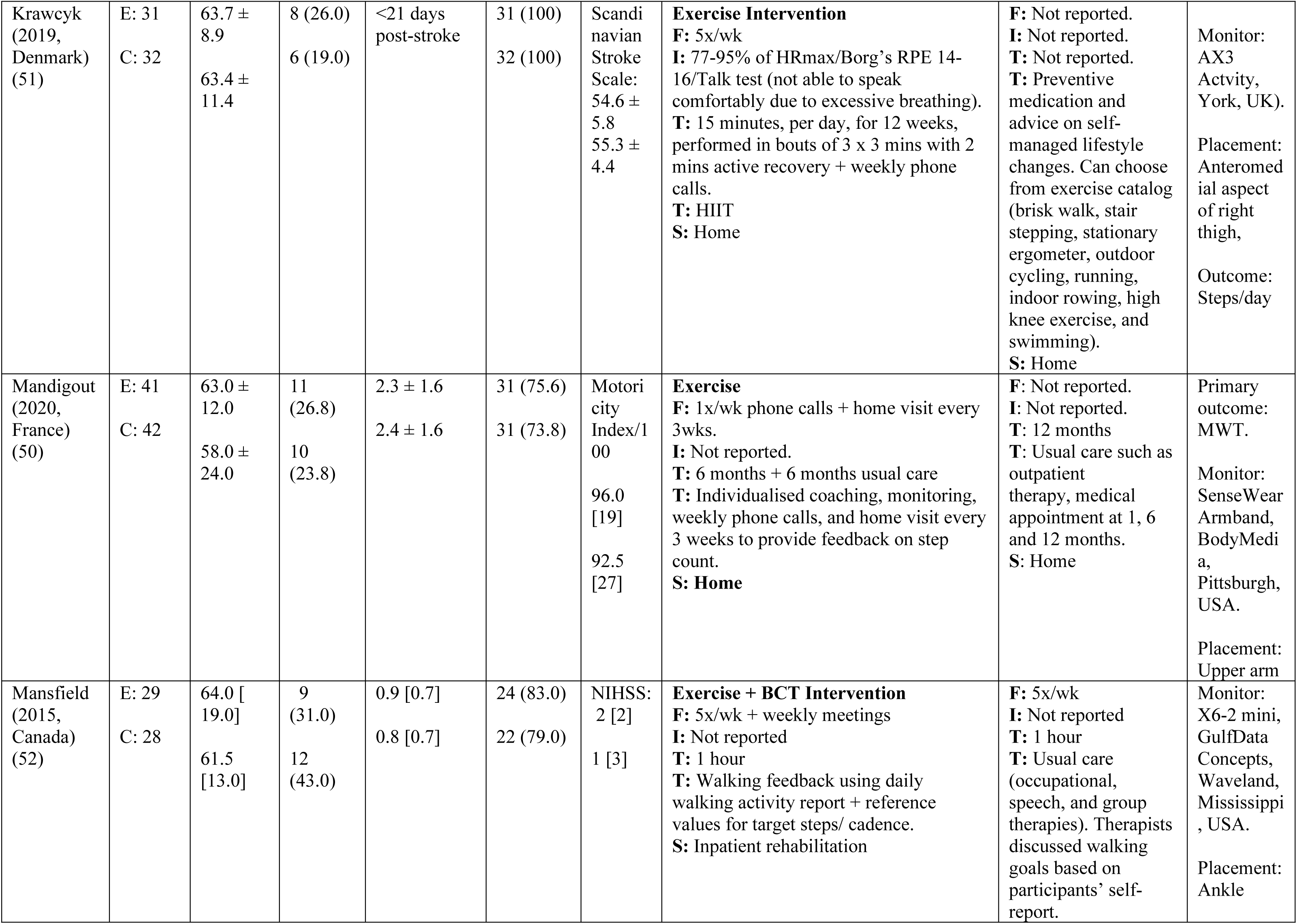
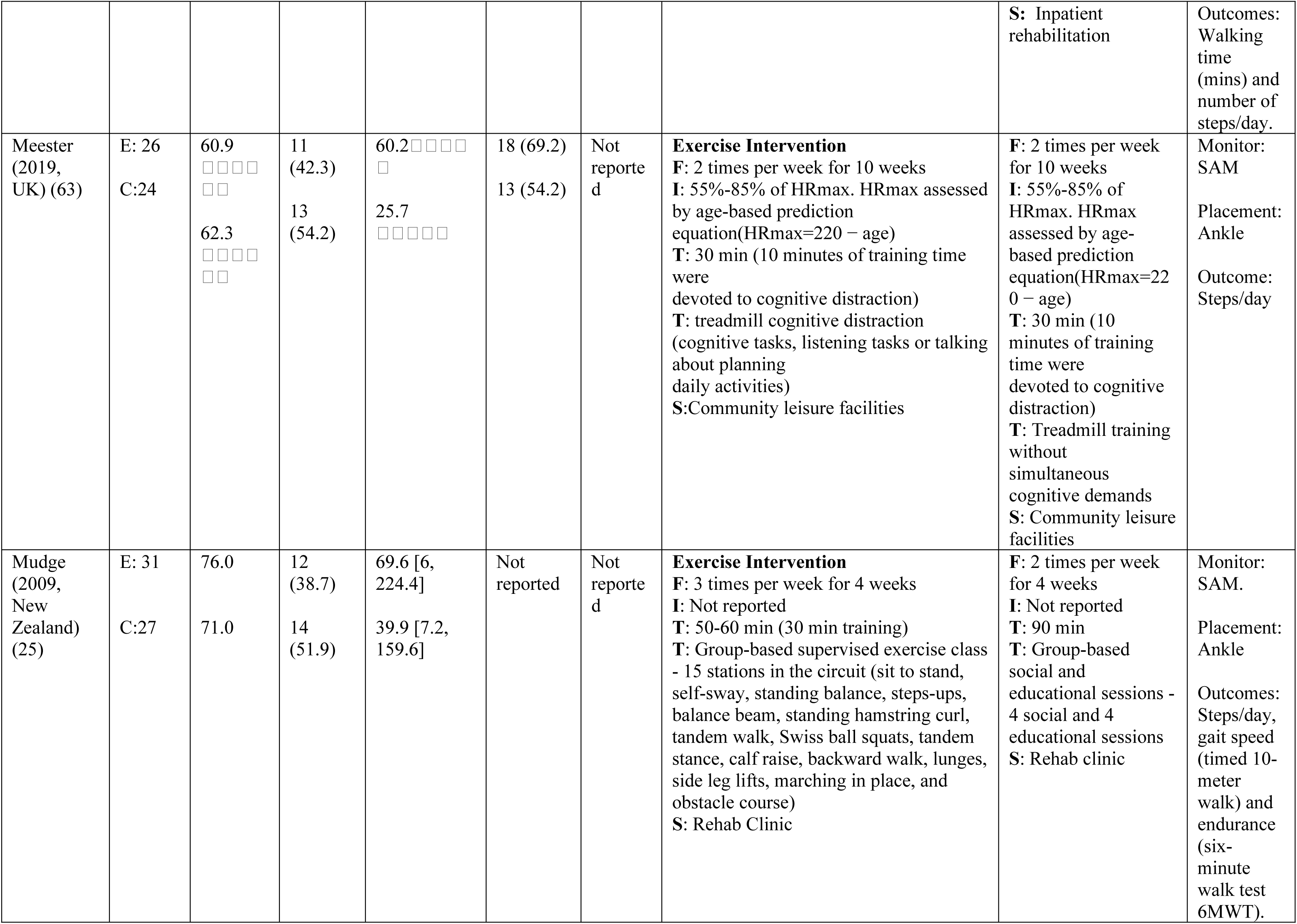
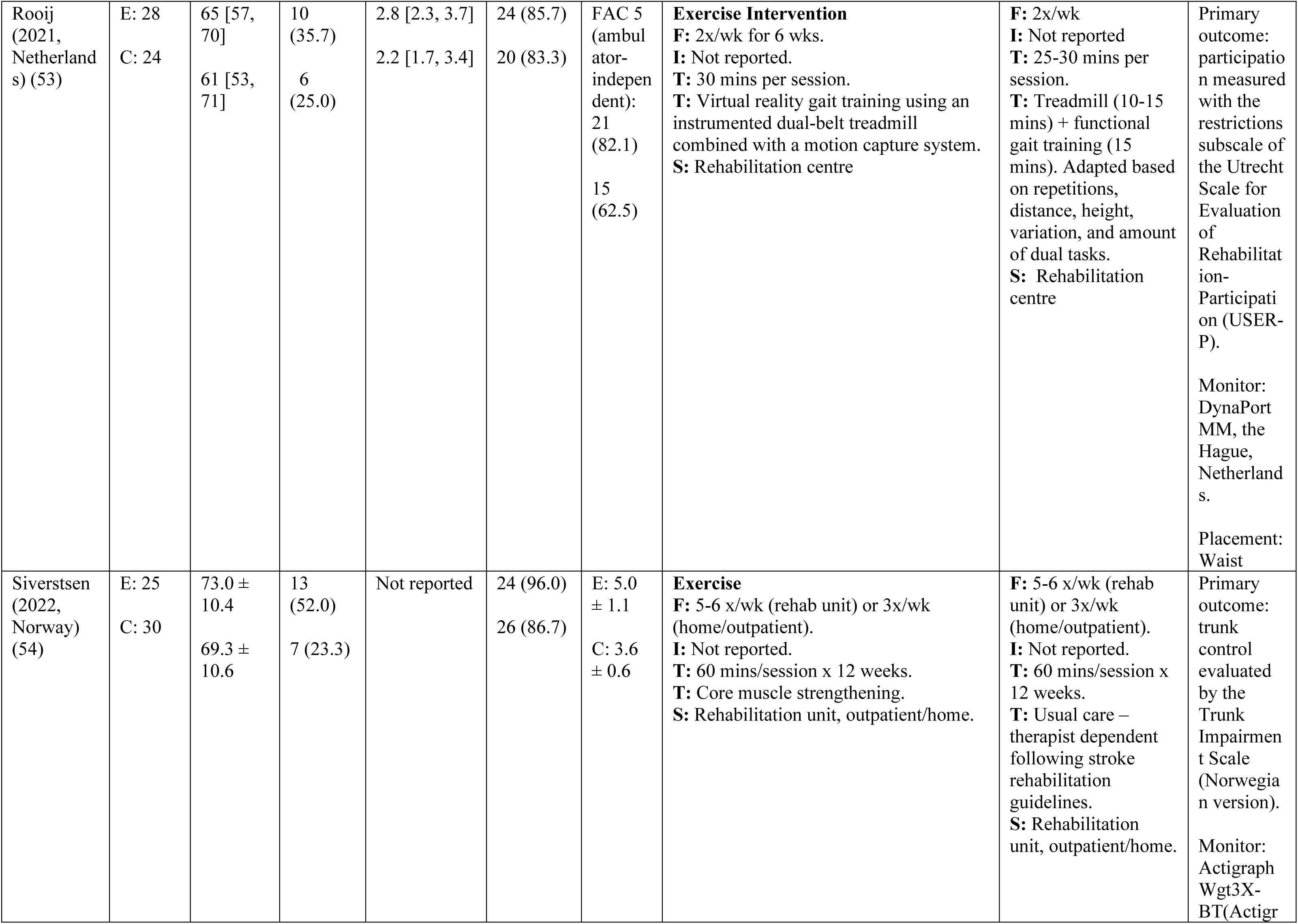
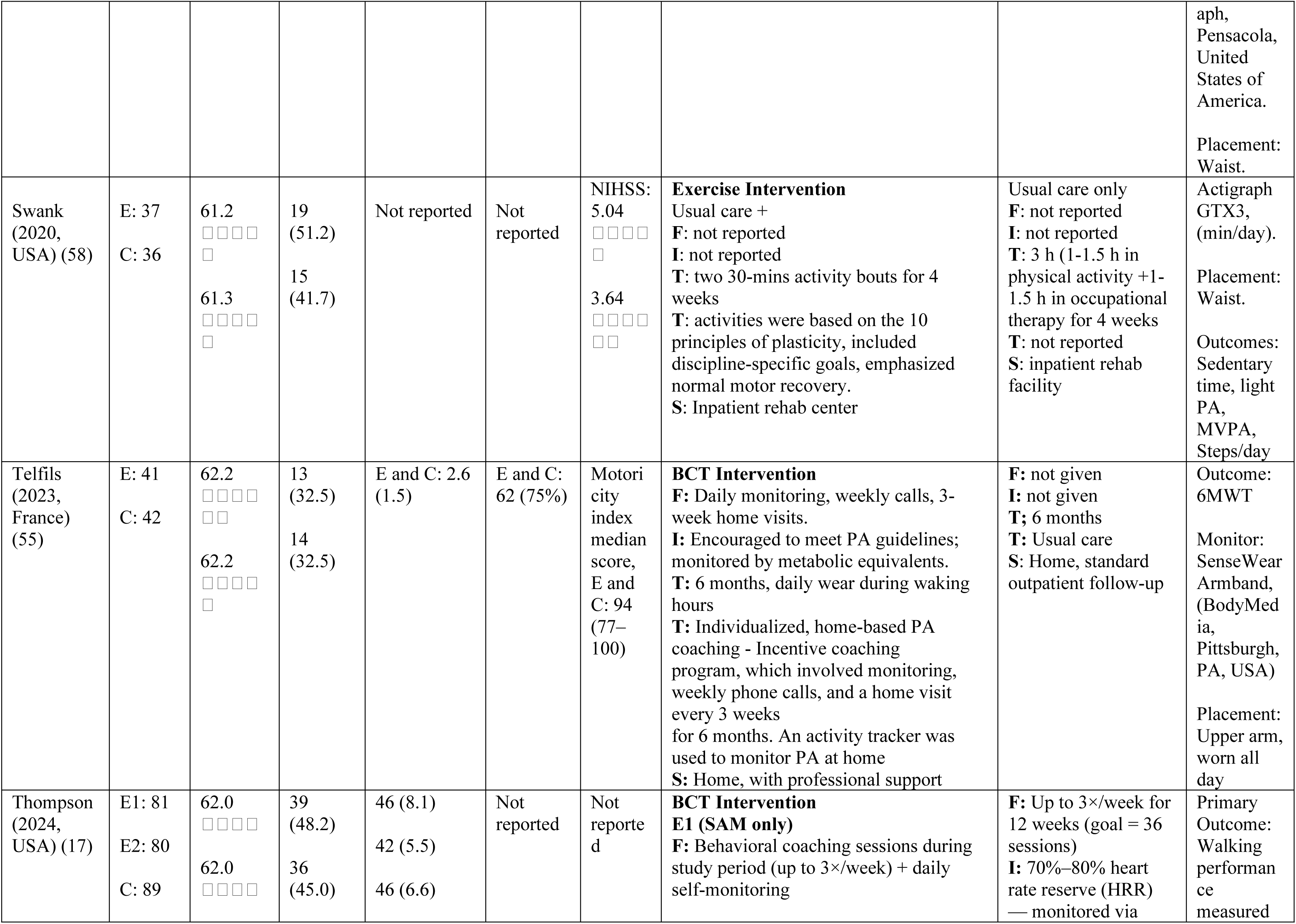
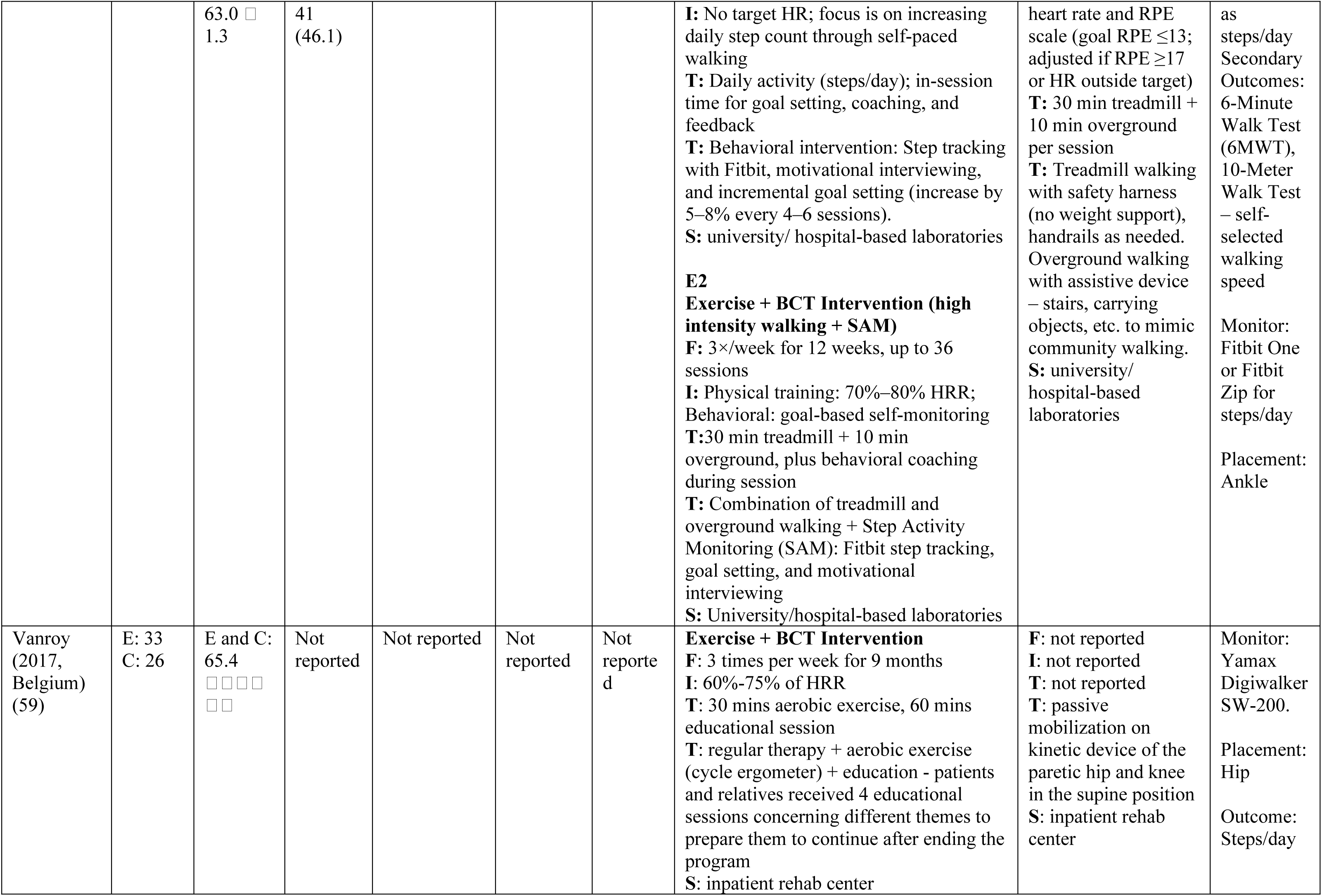
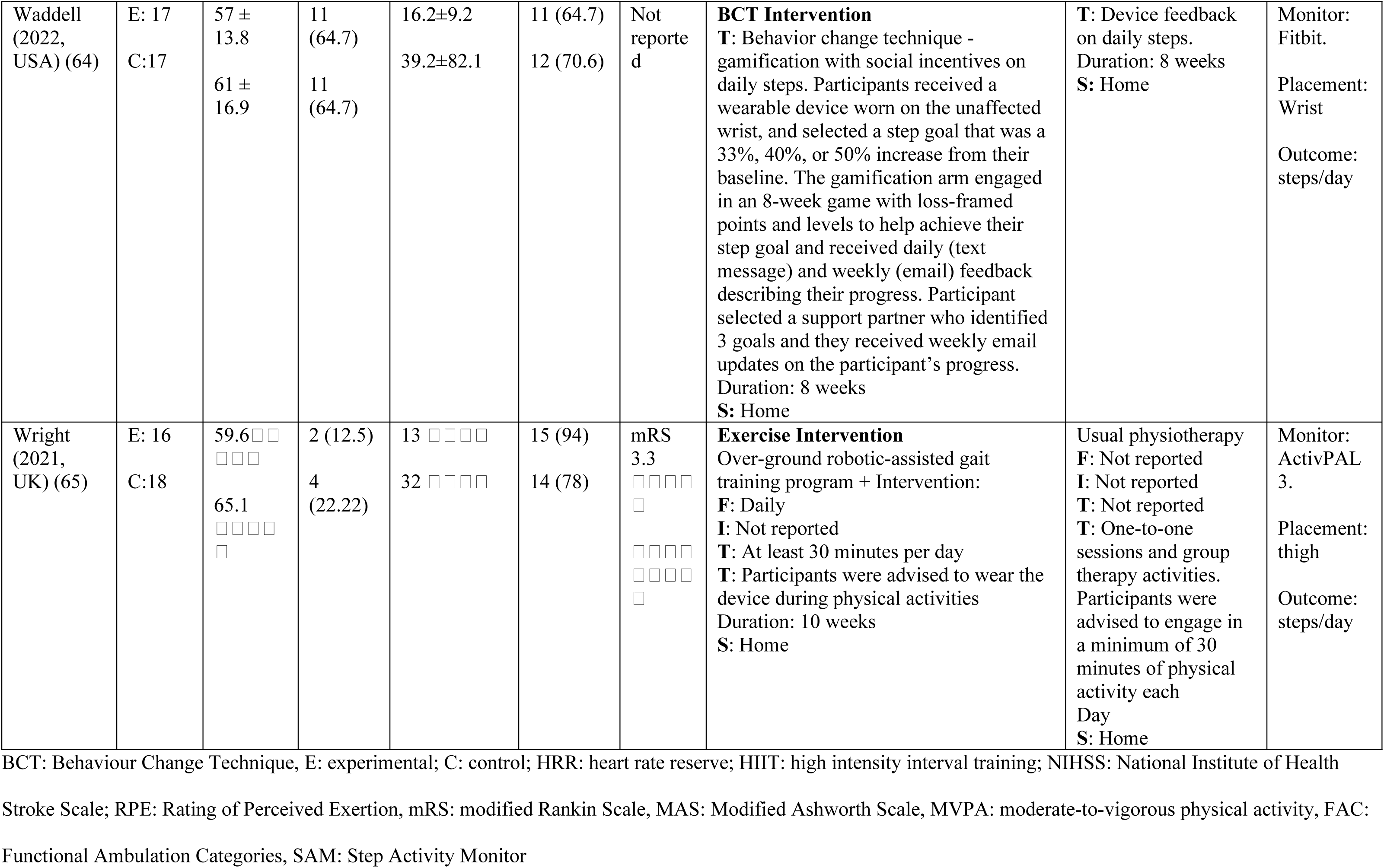
Characteristics of All Included Studies.

The mean age of participants in the experimental group ranged from 48 to 73.0 years, while for the control group, it ranged from 48 to 71.8 years. Fifteen studies (40–42,44,45,47,49–55,58,59) were on acute/subacute stroke phases, and 13 others were in the chronic stroke phase (17,25,43,46,48,56,57,60–65). Time since stroke was reported by 20 articles, with mean durations ranging from 0.12 to 60.2 months for the experimental group and 0.12 to 69.6 months for the control group. Ischemic stroke was the most prevalent type, affecting 64.7% to 100% of participants in the experimental group and 54.2% to 100% in the control group. Stroke severity, measured by the National Institutes of Health Stroke Scale (NIHSS) mean score, ranged from 0.9 to 6 for the intervention group and from 0.8 to 6 for the control group.

### Description of Interventions

Of the 28 articles qualitatively analyzed, 23 (17,25,40–43,45–47,49,51,52,54,56–65) reported performance measures, primarily focusing on steps per day (Table 2). Additionally, 18 studies (17,40–43,45–48,50,52–55,60–62,65) reported capacity outcomes, including comfortable gait speed, fastest gait speed, and/or the 6-Minute Walk Test (6MWT) (Table 3). The interventions employed across these studies generally fell into three main categories:

**Table 2.**
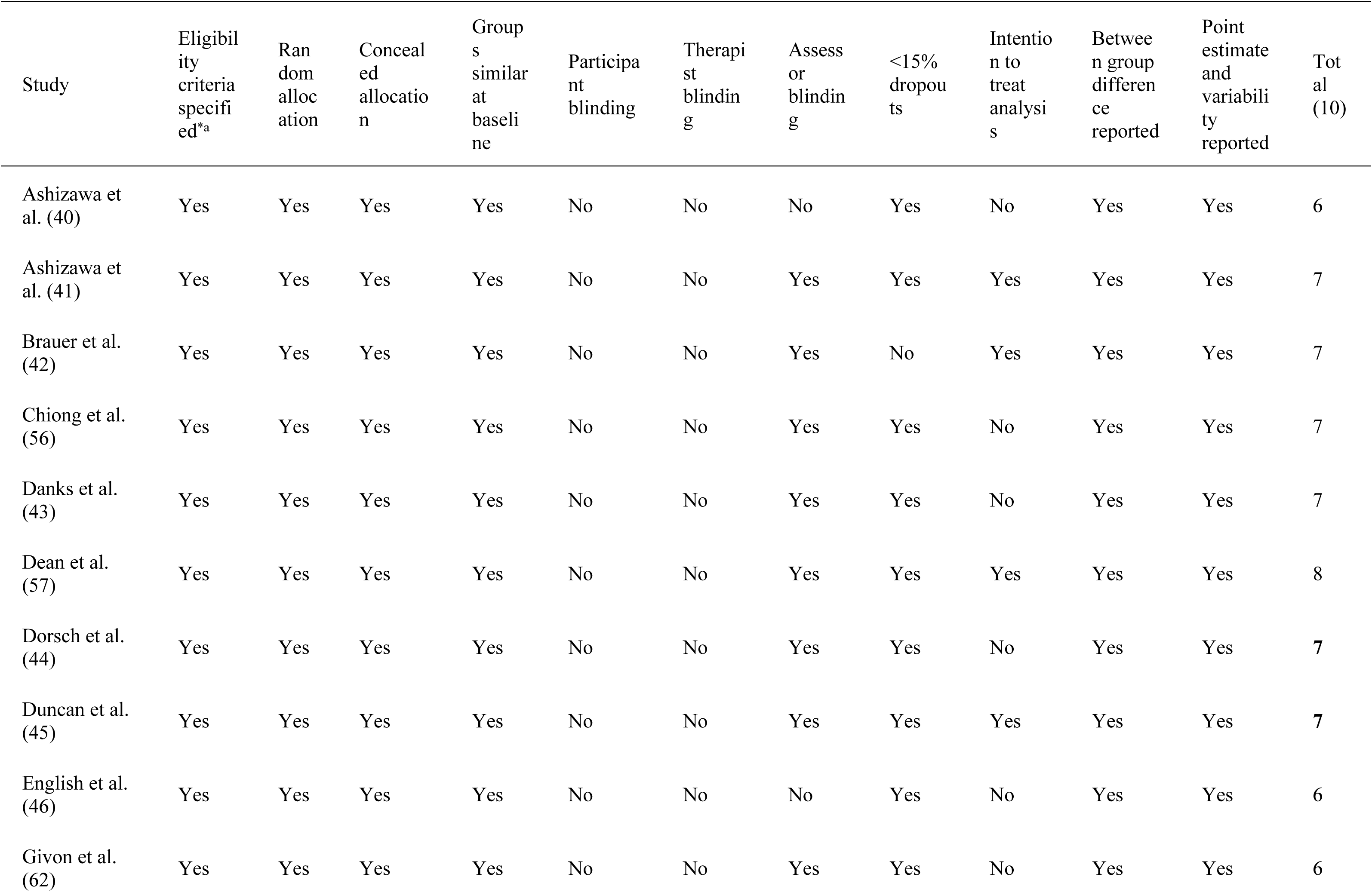
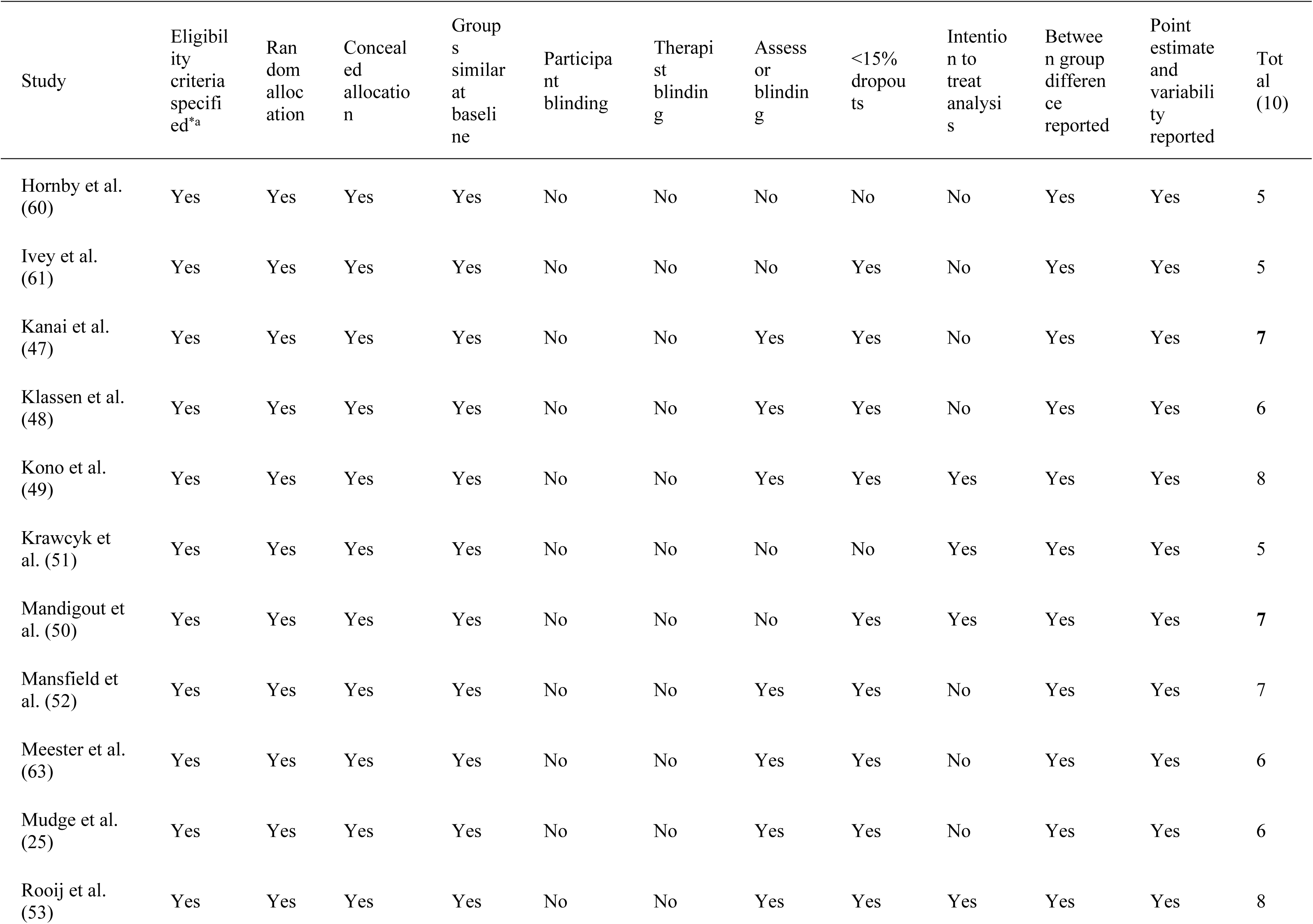
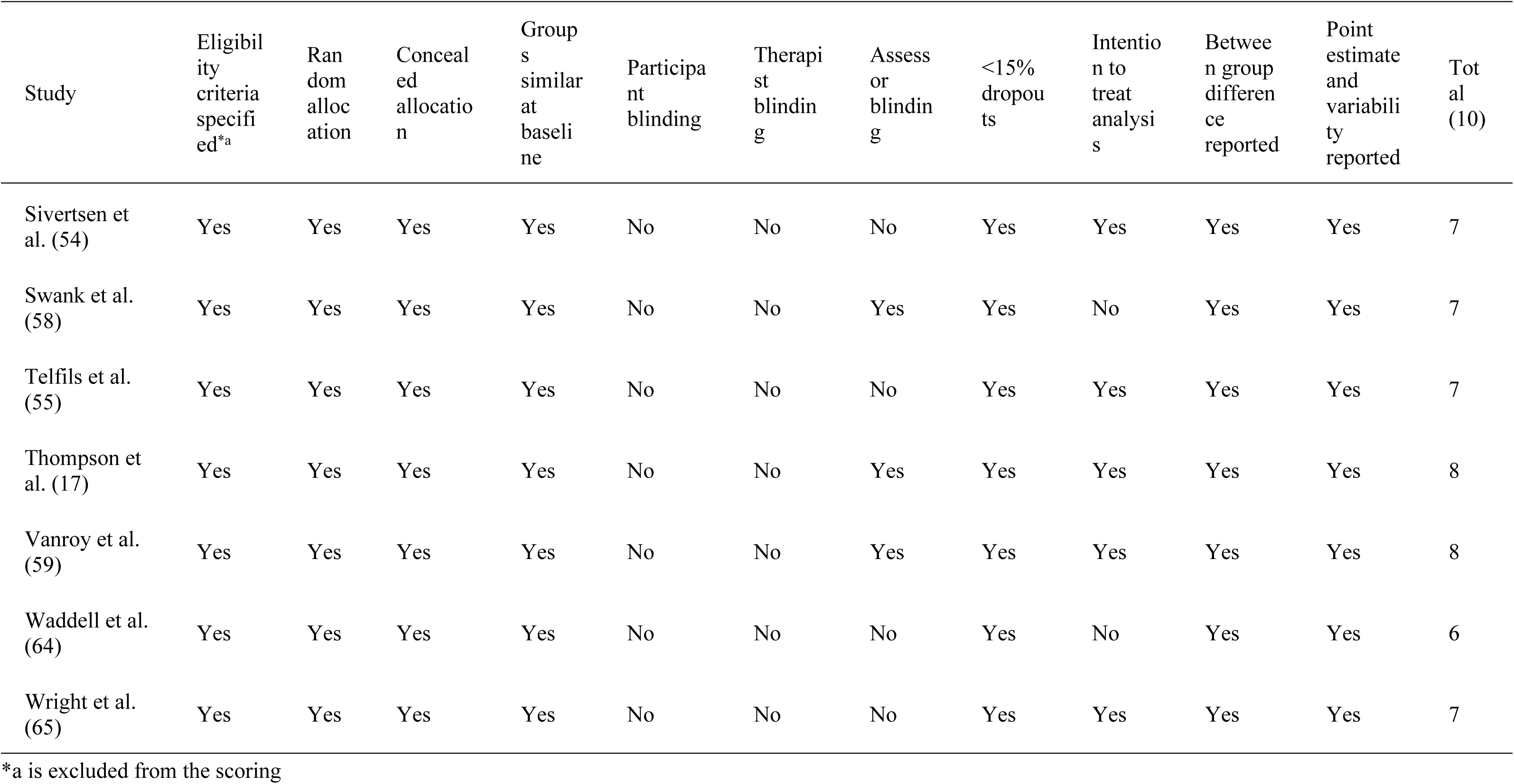
PEDro Scale Quality Assessment of Studies.

**Table 3:**
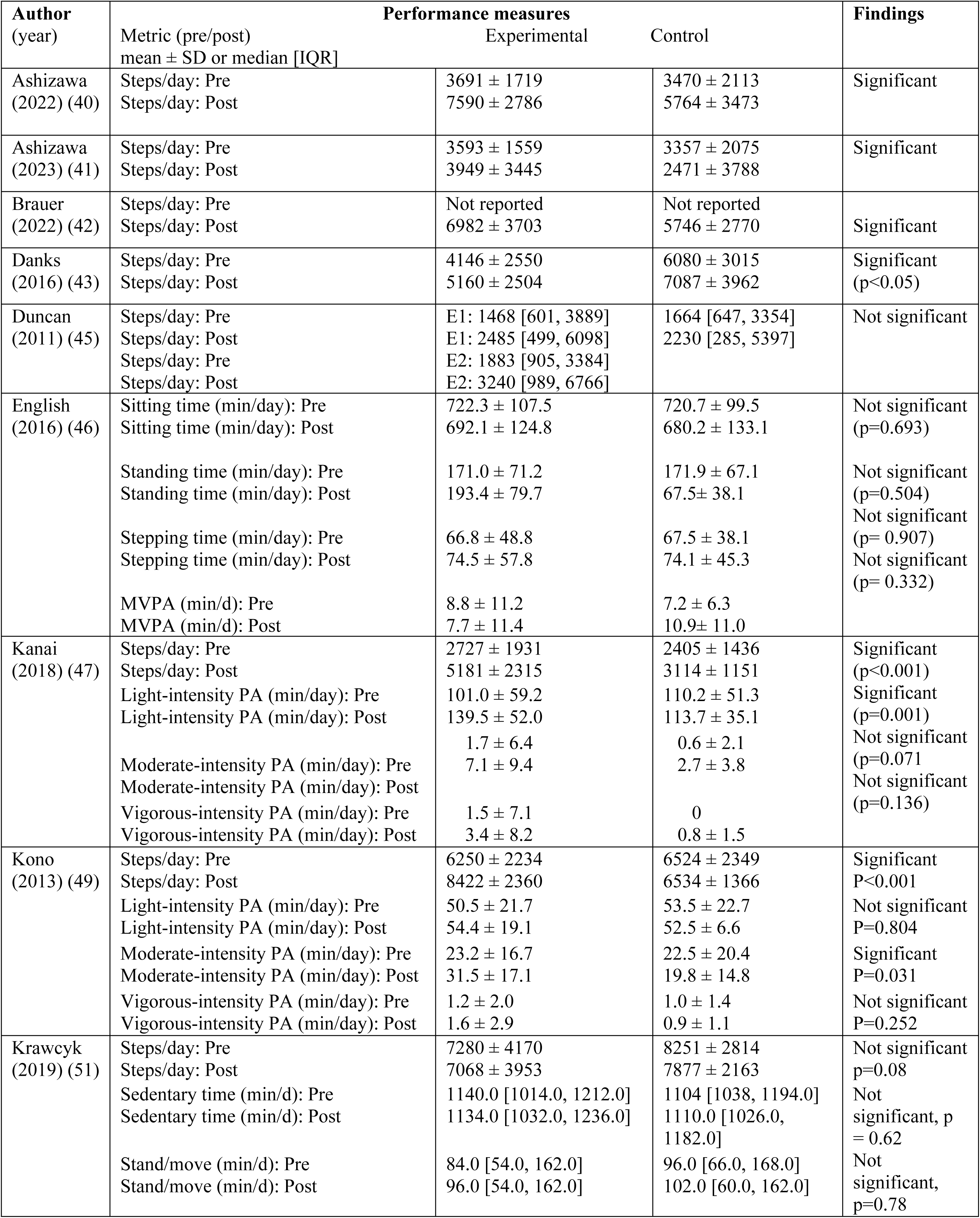
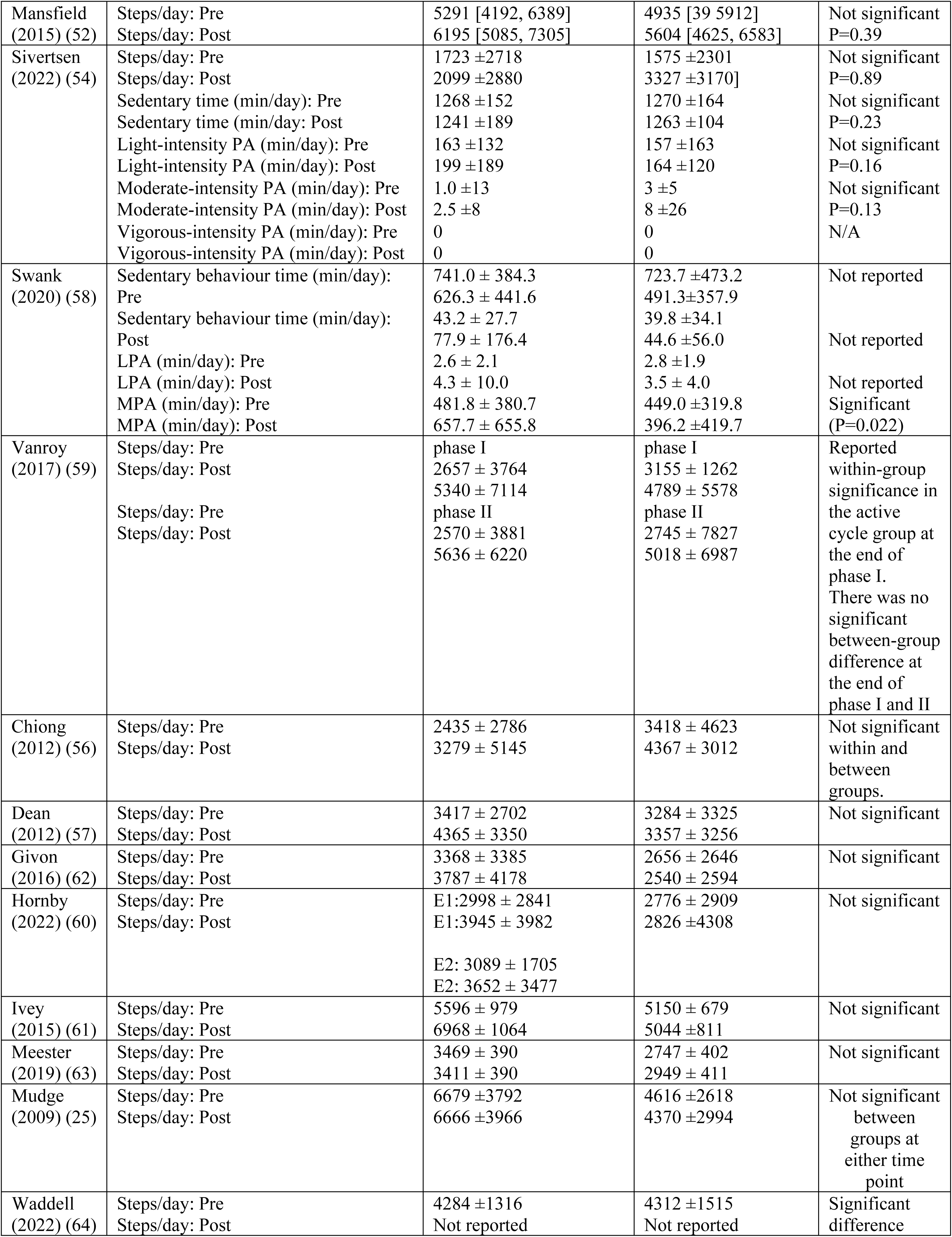
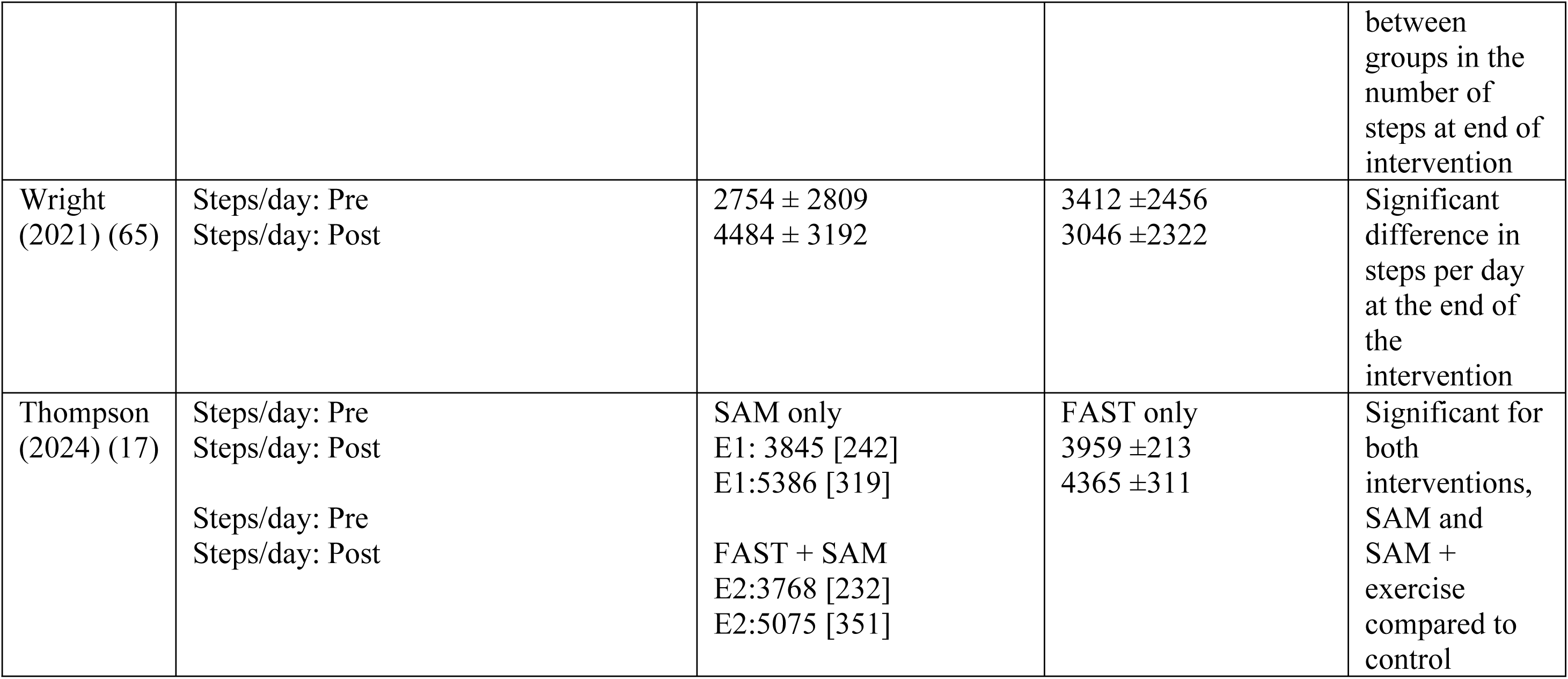
Performance measures of daily activity behavior.

### BCT Only

Seven studies reported interventions solely focused on BCTs. These interventions (alongside their BCT taxonomy) reported encompassed various strategies such as guidance/education and monitoring of physical activities with encouragement and praise (BCT 4.1 Instructions on how to perform the behaviour), goal-directed counselling (BCT 1.1 Goal setting - behaviour), self monitoring of SB and step counting (BCT 2.2 Self-monitoring of behaviours), encouragement and feedback on SB (BCT 2.2 Feedback on behaviour) (40,41); gamification incorporating social incentives to increase daily steps (BCT 1.1 Goal setting – behaviour (64); Education (BCT 5.1 Information about health Consequences), self-monitoring (BCT 2.2 self monitoring of behaviour); Goal setting (BCT 1.1 Goal setting - behavior) and coaching incentives (55); Goal directed counseling (BCT 1.1 Goal setting - behaviour) (46); Self-monitoring of behaviour and step counting (BCT 2.2 Self-monitoring of behaviour), (17); and augmented outcomes (BCT 2.7 feedback on outcome behaviour) (44).

- **Exercise Only:** Fifteen studies exclusively focused on exercise interventions. These interventions included high-intensity interval training (51), high-intensity treadmill training (60,61), treadmill training with concurrent cognitive demands (63), group circuit-based training (25), stretching exercises and orthotic device (57), overground locomotor training (45), virtual reality gait training (53), core muscle strengthening (I-CoreDIST) intervention (54), patient-directed activity programs (58), overground robotic-assisted gait training (65), progressive supervised walking exercise (48,50), group-based supervised exercise (25), and task-related training with progressive balance and strengthening exercises (57).
- **Combined BCT and Exercise:** Six studies (17,42,43,47,49,52) reported interventions that combined both BCTs and exercise. Brauer et al. (42) incorporated treadmill training with self-management strategies, including education (BCT 4.1), goal setting (BCT 1.1), self-monitoring of behaviour (BCT 2.3), and action planning (BCT 1.4). Other combined interventions include treadmill walking with behavioural goal setting (BCT 1.1) and self-monitoring (BCT 2.3) (17); a supervised rehabilitation program with accelerometer-based feedback and goal setting (BCT 1.1) (47); a mix of home-based and centre-based exercise, along with lifestyle advice and counselling (BCT 4.1 – instruction on how to perform the behaviour) (49); physiotherapy combined with accelerometer-based behavioural feedback (BCT 2.2) (52); regular therapy involving aerobic exercise and education (BCT 4.1) by Vanroy et al. (42); and treadmill and overground walking training with step activity monitoring (BCT 2.3 – self-monitoring of behaviour) (43).

### Activity Monitors

Across the studies, 26 mentions of activity monitors were identified. The StepWatch Activity Monitor was the most frequently used, cited 5 times (19.23%) (19.23%)(17,25,60,61,63) of all monitor mentions. This device was placed on the ankle of the paretic limb, indicating its suitability for measuring lower limb activity in stroke populations. Three other monitors were each mentioned 3 times (11.54%): ActivPAL3 (PAL Technologies) (42,46,65), typically worn on the thigh to assess 24-hour movement behaviour and transitions in posture; Actigraph (GT3+, Wgt3X-BT, GTX3): (46,54,58), commonly placed on the waist to measure whole-body movement and energy expenditure; and Fitbit (One, Zip) (47,48,64), with one mention specifying the nonparetic ankle and others unspecified, suggesting flexible placement. Two other devices were each mentioned twice (7.69%): the Active Style Pro HJA-750C (Omron)(40,41), worn on the wrist, and Gulf Coast Data (44,52) Concepts (X6-2 mini) placed on the ankle Devices mentioned once (3.85% each) included the Kenz Lifecorder, AX3 (York) (49), DynaPort MM (53), SenseWear Armband (BodyMedia) (55), a generic Pedometer (56), Digimax pedometer (57), Yamax Digiwalker SW-200 (59), and Actical Minimiter Co (62). Their placements varied, with the AX3 on the anteromedial aspect of the right thigh, DynaPort MM on the waist, and SenseWear Armband on the upper arm.

### Risk of Bias

The PEDro scores ranged from 5 to 8 points (Table 2). No study achieved the maximum score of 10, as blinding of participants and treatment providers was not feasible for these types of interventions. Of the 28 studies assessed, 25 (89.3%) scored 6 or higher and were therefore considered high quality, while only 3 studies (10.7%) scored 5 and were rated as moderate quality (51,60,61).

### Narrative Syntheses

We included narrative synthesis of findings where outcomes were reported in single studies or where the outcomes reported were too diverse or different measurements were used that did not allow data to be pooled in a meta-analysis.

#### Performance Outcomes

Steps per day was the most reported performance outcome in 23 out of 28 included studies (17,25,40–43,45–47,49,51,52,54,56–65) (see Table 3), with 9 studies reporting statistically significant results at post-intervention (17,40–42,47,49,58,64,65). Conversely, 11 studies reported no significant findings for steps per day (25,43,45,51,52,54,57,60–63). Vanroy et al. (59) reported within-group significance in an active cycle group at the end of one phase but no significant between-group difference across both phases of their intervention.

Beyond daily step count, Table 3 also includes findings for other performance-related outcomes, such as sedentary time (46,51,54,58), stepping time (46,51), as well as PA intensity gradations such as light, moderate, and vigorous. Three (46,51,54) of the 4 studies on sedentary time reported no significant findings for sedentary time. For stepping time, English et al. (46) reported a significant within-group effect, but not between-group effect. Regarding light-intensity PA (LPA), 2 studies (49,54) reported no significant effects, while Kanai et al. (47) found a significant effect. Four studies reported non significant findings for moderate-intensity PA (MPA) (46,47,49,54) and 3 studies (47,49,54) reported non significant findings for vigorous-intensity PA (VPA).

In summary, a notable portion of the included studies demonstrated improvements in daily step count, with some interventions also showing significant effects on stepping time and certain intensities of PA, while other metrics like sedentary time and light-intensity activity often did not reach statistical significance at the individual study level as reported in Table 3.

#### Capacity Outcomes

Table 4 summarizes the physical capacity outcomes reported across 18 studies (17,25,40–43,45–48,50,52–55,60–62,65). The primary measures include the 6-Minute Walk Test (6MWT) for endurance, as well as comfortable (or self-selected) and fastest gait speeds.

**Table 4:** Physical capacity measures of walking speed and endurance.

| S/N | Author<br>(year) | Capacity measures |  |  | Findings |
| --- | --- | --- | --- | --- | --- |
|  |  | Time point | Experimental | Control |  |
| 1 | Ashizawa<br>(2022) (40) | 6MWT (m): Pre<br>6MWT (m): Post<br>Comfortable gait speed (m/s): Pre<br>Comfortable gait speed (m/s):<br>Post | 489.5 ± 87.1<br>Not reported<br>1.5 ± 0.3<br>Not reported | 477.8 ± 100.9<br>Not reported<br>1.4 ± 0.3<br>Not reported | Not significant<br>at baseline<br>(p=0.64)<br>Not significant<br>at baseline<br>(p=0.11) |
| 2 | Ashizawa<br>(2023) (41) | 6MWT (m): Pre<br>6MWT (m): Post<br>Comfortable gait speed (m/s): Pre<br>Comfortable gait speed (m/s):<br>Post | 490.8 ± 97.9<br>Not reported<br>1.5 ± 0.3<br>Not reported | 481.0 ± 97.5<br>Not reported<br>1.4 ± 0.3<br>Not reported | Not significant<br>(p=0.63) at<br>baseline.<br>Not significant<br>(p=0.71) at<br>baseline |
| 3 | Brauer (2022)<br>(42) | Comfortable gait speed (m/s): Pre<br>Comfortable gait speed (m/s):<br>Post<br>Fastest gait speed (m/s): Pre<br>Fastest gait speed (m/s): Post<br>6MWT (m): Pre<br>6MWT (m): Post | 0.94 ± 0.30<br>1.19 ± 0.30<br>1.22 ± 0.40<br>1.51 ± 0.41<br>333.0 ± 117.0<br>423.0 ± 116.0 | 0.88 ± 0.29<br>1.05 ± 0.26<br>1.12 ± 0.38<br>1.31 ± 0.38<br>304.0 ± 111.0<br>370.0 ± 101.0 | Not significant<br>for<br>comfortable<br>gait speed,<br>fastest gait<br>speed and<br>6MWT |
| 4 | Danks<br>(2016) (43) | Comfortable gait speed (m/s): Pre<br>Comfortable gait speed (m/s):<br>Post<br>Fastest gait speed (m/s): Pre<br>Fastest gait speed (m/s): Post | 0.57 ± 0.36<br>0.68 ± 0.37<br>0.67 ± 0.50<br>0.88 ± 0.54 | 0.68 ± 0.41<br>0.76 ± 0.44<br>0.84 ± 0.54<br>0.94 ± 0.60 | Significant for<br>6MWT,<br>comfortable<br>and fastest gait<br>speed (p<0.05) |
| 5 | Duncan (2011)<br>(45) | 6MWT (m): Pre<br>6MWT (m): Post<br>6MWT (m): Pre<br>6MWT (m): Post<br>Comfortable gait speed (m/s): Pre<br>Comfortable gait speed (m/s):<br>Post<br>Comfortable gait speed (m/s): Pre<br>Comfortable gait speed (m/s):<br>Post | E1: 124.1 ± 77.5<br>E1: 205.9 ± 99.9<br>E2: 126.3 ± 75.0<br>E2: 202.2 ± 100.5<br>E1: 0.37 ± 0.22<br>E1: 0.62 ± 0.30<br>E2: 0.39 ± 0.22<br>E2: 0.62 ± 0.30 | 125.7 ± 81.8<br>166.7 ± 94.0<br>0.38 ± 0.23<br>0.51 ± 0.27 | Not significant<br>for 6MWT<br>across the<br>groups<br>(p=0.97)<br>Not significant<br>for<br>comfortable<br>gait speed<br>across the<br>groups<br>(p=0.75) |
| 6 | English (2016)<br>(46) | Comfortable gait speed (m/s): Pre<br>Comfortable gait speed (m/s):<br>Post | 0.80 ± 0.36<br>Not reported | 0.82 ± 0.51<br>Not reported | Not reported |
| 7 | Kanai (2018) (47) | Comfortable gait speed (m/s): Pre<br>Comfortable gait speed (m/s):<br>Post | 1.0 ± 0.2<br>Not reported | 1.0 ± 0.3<br>Not reported | Not reported |
| 8 | Klassen (2020)<br>(48) | 6MWT (m): Pre<br>6MWT (m): Post | E1: 129.0 ± 97.3<br>E1: 246 ± 138.0 | 129.0 ± 77.6<br>307.0 ± 138.0 | Significant for<br>6MWT and<br>comfortable<br>gait speed for<br>both DOSE |
|  |  | 6MWT (m): Pre<br>6MWT (m): Post<br>Comfortable gait speed (m/s): Pre<br>Comfortable gait speed (m/s): Post<br>Comfortable gait speed (m/s): Pre<br>Comfortable gait speed (m/s): Post | E2: 138.0 ± 95.5<br>E2: 315.0 ± 142.0<br>E1: 0.44 ± 0.25<br>E1: 0.90 ± 0.31<br>E2: 0.42 ± 0.25<br>E2: 0.97 ± 0.40 | 0.39 ± 0.22<br>0.74 ± 0.37 | interventions compared to control |
| 9 | Mandigout (2021) (50) | 6MWT (m): Pre<br>6MWT (m): Post | 369.0 ± 162.0<br>418.0 ± 122.2 | 342.0 ± 220.0<br>389.0 ± 139.3 | Not significant between group findings at any time point |
| 10 | Mansfield (2015) (52) | Comfortable gait speed (m/s): Pre<br>Comfortable gait speed (m/s): Post | 0.83 ± 0.32<br>1.02 ± 0.28 | 0.80 ± 0.45<br>0.85 ± 0.43 | Significant P=0.025 |
| 11 | Rooji (2021) (53) | 6MWT (m): Pre<br>6MWT (m): Post | 359.5 ± 124.3<br>408.2 ± 125.3 | 357.8 ± 104.1<br>428.5 ± 110.4 | Not significant |
| 12 | Sivertsen (2022) (54) | Comfortable gait speed (m/s): Pre<br>Comfortable gait speed (m/s): Post<br>Fastest gait speed (m/s): Pre<br>Fastest gait speed (m/s): Post | 0.72 ± 0.47<br>0.94 ± 0.30<br>1.05 ± 0.70<br>1.28 ± 0.44 | 0.80 ± 0.44<br>1.05 ± 0.26<br>1.10 ± 0.57<br>1.46 ± 0.47 | Not significant (between-group) P = 0.23<br>Not significant (between-group) P = 0.11 |
| 13 | Hornby (2022) (60) | Comfort. gait speed (m/s): Pre<br>Comfort. gait speed (m/s): Post<br>Comfort. gait speed (m/s): Pre<br>Comfort. gait speed (m/s): Post<br>Fastest gait speed (m/s): Pre<br>Fastest gait speed (m/s): Post<br>Fastest gait speed (m/s): Pre<br>Fastest gait speed (m/s): Post<br>6MWT (m): Pre<br>6MWT (m): Post<br>6MWT (m): Pre<br>6MWT (m): Pos | E1: 0.54 ± 0.30<br>E1: 0.16 ± 0.13<br>E2: 0.51 ± 0.33<br>E2: 0.22 ± 0.19<br>E1: 0.73 ± 0.41<br>E1: 0.24 ± 0.19<br>E2: 0.68 ± 0.40<br>E2: 0.35 ± 0.24<br>E1: 212.0 ± 138.0<br>E1: 294.0 ± 149.7<br>E2: 197.0 ± 132.8<br>E2: 293.0 ± 152.8 | 0.48 ± 0.30<br>0.50 ± 0.30<br>0.60 ± 0.40<br>0.35 ± 0.13<br>197.0 ± 140.0<br>231.0 ± 145.0 | Significant for comfortable gait speed, Fastest gait speed and 6MWT for both HIIT interventions compared to control |
| 14 | Ivey (2015) (61) | 6MWT (m): Pre<br>6MWT (m): Post | 780 ± 105<br>964 ± 131 | 564 ± 73<br>668 ± 76 | Not significant (between-group) P = 0.11 |
| 15 | Givon (2016) (62) | Gait speed (m/s): Pre<br>Gait speed (m/s): Post | 0.9 (0.4)<br>1.0 (0.4) | 0.7 (0.3)<br>0.8 (0.4) | Significant (p=0.02) |
| 16 | Mudge (2009) (25) | Gait speed (m/s): Pre<br>Gait speed (m/s): Post<br>6MWT (m): Pre | 0.76 ± 0.30<br>0.79 ± 0.28<br>263 ± 110<br>282 ± 117 | 0.62 ± 0.27<br>0.63 ± 0.25<br>201 ± 99<br>200 ± 99 | Significant for gait speed (between-group) (P=0.038) |
|  |  | 6MWT (m): Post |  |  | Significant for 6MWT (between-group) P = 0.030 |
| 17 | Wright (2021) (65) | 6MWT (m): Pre<br>6MWT (m): Post | 135 ± 81<br>158 ± 93 | 122 ± 92<br>119 ± 84 | No significant difference |
| 18 | Telfils (2022) (55) | 6MWT (m): Pre<br>6MWT (m): Post | 361.9 ± 148.4<br>430.8 ± 145.1 | 379.9 ± 147.8<br>391.6 ± 152.5 | Significant (p=0.01) |
| 19 | Thompson (2024) (17) | 6MWT (m): Pre<br>6MWT (m): Post<br>6MWT (m): Pre<br>6MWT (m): Post<br>Self-selected gait speed (m/s): Pre<br>Self-selected gait speed (m/s): Post<br>Self-selected gait speed (m/s): Pre<br>Self-selected gait speed (m/s): Post | E1: 279 ± 12<br>E1: 304 ± 13<br>E2: 283 ± 14<br>E2: 323 ± 15<br>SAM only:<br>E1: 0.68 ± 0.02<br>E1: 0.79 ± 0.03<br>FAST + SAM:<br>E2: 0.68 ± 0.03<br>E2: 0.82 ± 0.03 | 296 ± 12<br>338 ± 13<br>FAST only:<br>0.71 ± 0.02<br>0.84 ± 0.03 | Significant within-group effect for 6MWT and self-selected gait speed for both interventions compared to the control. Not significant for between-groups for 6MWT and self-selected gait speed. |

Notes: 6MWT- 6 Minute Walk test, FAST - High-intensity walking intervention, SAM- Step Activity Monitoring, HIIT - high intensity interval training

**Table 5:** Standardised Mean Differences (SMD) and their 95% Confidence Intervals (CI) for studies included in the meta-analysis.

| Study | Intervention | Control | Stroke Stage | SMD (steps) (95% CI) | SMD (comfortable gait speed) (95% CI) | SMD (fast gait speed) (95% CI) | SMD (6MWT) (95% CI) |
| --- | --- | --- | --- | --- | --- | --- | --- |
| Thompson, 2024a (17) | BCT | Exercise | Chronic | 0.35 (0.05, 0.65) | NA | NA | -0.28 (-0.58, 0.02) |
| Thompson, 2024b (17) | BCT + Exercise | Exercise | Chronic | 0.23 (-0.07, 0.53) | -0.07 (-0.37, 0.23) | NA | -0.12 (-0.42, 0.18) |
| Ashizawa, 2023 (41) | BCT (Education on reducing SB) | Exercise (Education on increasing PA) | Acute | 0.40 (-0.02, 0.83) | NA | NA | NA |
| Ashizawa, 2022 (40) | BCT (Education on reducing SB) | Exercise (Education on increasing PA) | Acute | 0.57 (0.07, 1.08) | NA | NA | NA |
| Sivertsen, 2022 (54) | Exercise (I-CoreDIST intervention) | Usual care | Subacute | -0.40 (-0.90, 0.11) | -0.39 (-0.89, 0.12) | -0.44 (-0.95, 0.06) | NA |
| Hornby, 2022a (60) | Exercise (High-intensity training in variable context) | Exercise (Low-intensity variable stepping training) | Chronic | 0.38 (-0.24, 1.00) | 0.65 (0.02, 1.28) | 0.73 (0.09, 1.36) | 0.71 (0.07, 1.34) |
| Hornby, 2022b (60) | Exercise (High-intensity forward training with minimal variability) | Exercise (Low-intensity variable stepping training) | Chronic | 0.34 (-0.28, 0.96) | 0.72 (0.08, 1.35) | 0.84 (0.20, 1.48) | 0.66 (0.03, 1.29) |
| Wright, 2021 (65) | Exercise + Therapeutic technology | Usual care | Chronic | 0.51 (-0.16, 1.18) | NA | NA | 0.43 (-0.25, 1.11) |
| Swank, 2020 (58) | Exercise (patient-directed) | Usual care | Subacute | 0.47 (0.01, 0.93) | NA | NA | NA |
| Krawczyk, 2019 (51) | Exercise (High intensity interval training) | Usual care | Subacute (12 weeks) | -0.25 (-0.74, 0.24) | NA | NA | NA |
| Meester, 2019 (63) | Exercise (Treadmill training with simultaneous cognitive demand) | Exercise (Treadmill training without cognitive demands) | Chronic | 0.23 (-0.32, 0.78) | NA | NA | NA |
| Kanai, 2018 (47) | Exercise + BCT | Supervised rehabilitation program | Acute | 1.12 (0.56, 1.68) | NA | NA | NA |
| Danks, 2016 (43) | BCT + Exercise | Exercise (Aerobic) | Chronic | -0.56 (-1.31, 0.19) | -0.19 (-0.83, 0.44) | -0.15 (-0.78, 0.48) | 0.00 (-0.73, 0.73) |
| Givon, 2016 (62) | Exercise (Whole body video game exercise) | Usual care | Chronic | 0.35 (-0.22, 0.92) | 0.49 (-0.08, 1.06) | 0.67 (0.09, 1.25) | NA |
| Mansfield, 2015 (52) | BCT + Exercise | Usual care | Subacute | 0.21 (-0.29, 0.72) | 0.44 (-0.06, 0.95) | NA | NA |
| Chiong, 2012 (56) | Exercise and orthotic device | Exercise (Stretching) | Chronic | -0.22 (-1.39, 0.95) | NA | NA | NA |
| Dean, 2012 (57) | Exercise (designed to enhance mobility and PA) | Exercise (designed to enhance upper limb mobility and cognition) | Chronic | 0.30 (-0.02, 0.62) | NA | NA | NA |
| Mudge, 2009 (25) | Exercise (group-based supervised exercise class) | Group based social and educational sessions | Chronic | 0.64 (0.12, 1.16) | NA | NA | 0.77 (0.24, 1.29) |
| Brauer, 2022 (42) | BCT + Exercise | Usual gait training | Acute | 0.38 (-0.01, 0.76) | 0.50 (0.13, 0.86) | 0.62 (0.25, 0.99) | NA |
| Klassen, 2020a (48) | Exercise (DOSE 1 – 30 minutes moderate intensity $\geq 40\%$ progressing to $>60\%$ HRR) | Usual care | Early subacute (1-4 weeks) | NA | 0.46 (-0.09, 1.01) | NA | 0.47 (-0.09, 1.02) |
| Klassen, 2020b (48) | DOSE 2 - Higher intensity training (extra 1-hour session in addition to DOSE1) | Usual care | Early subacute (1-4 weeks) | NA | 0.59 (0.03, 1.15) | NA | 0.49 (-0.07, 1.04) |
| Duncan, 2011a (45) | Exercise (Locomotor training using body weight) | Exercise (Home-based) | Subacute | NA | 0.38 (0.14, 0.63) | NA | 0.40 (0.16, 0.65) |
| Duncan, 2011b (45) | Exercise | Exercise (Home-based) | Subacute | NA | 0.38 (0.14, 0.62) | NA | 0.36 (0.12, 0.60) |
| Telflis, 2023 (55) | BCT | Usual care | Subacute ($>6$ months) | NA | NA | NA | 0.26 (-0.17, 0.69) |
| Rooij, 2021 (53) | Exercise (Virtual reality gait training) | Exercise (Treadmill training) | Subacute | NA | NA | NA | -0.17 (-0.71, 0.37) |
| Kono, 2013 (49) | BCT + Exercise | Advise on lifestyle modification | Subacute | 0.79 (0.28, 1.30) | NA | NA | NA |
| Mandigout, 2020 (50) | Exercise | Usual care | Subacute | NA | NA | NA | 0.16 (-0.26, 0.52) |

For endurance, measured by the 6MWT, significant improvements were reported in several studies (25,43,48,55,60). Notably, Klassen et al. (48) found significant benefits for both DOSE interventions compared to control, and Hornby et al. (60) reported similar effects for both HIIT interventions. Mudge et al. (25) also observed significant between-group differences. In the PROWALK trial (17), both the SAM and SAM + exercise groups showed significant within-group improvements, although between-group differences were not statistically significant. Other studies reported only baseline 6MWT data without follow-up (40,41,46,47) while several reported no significant changes in walking endurance (42,45,50,61,65).

Comfortable or self-selected gait speed was assessed in 12 studies (17,40–43,45–48,52,54,60), with significant improvements reported in six of them (17,25,43,48,60,62). These findings highlight meaningful changes in walking speed under usual effort conditions across multiple intervention types. Fastest gait speed was less frequently reported. Danks et al. (43) found a significant improvement, whereas three other studies (42,54,60) did not.

### Meta-analyses results

#### Performance Outcomes

##### Meta-analysis of the effect of exercise-only interventions on daily steps

Meta-analysis of exercise-only interventions on daily steps (Figure 2A) included ten studies, (25,51,54,56–58,60,62,63,65) comprising 313 participants in the experimental groups and 310 in the control groups. Using a random effects model, the standardized mean difference (SMD) was 0.23 (95% CI: 0.03 to 0.44), indicating a statistically significant positive effect of exercise on step count with a moderate quality of evidence. Heterogeneity was moderate, with an I^2^ value of 36.57%, τ² = 0.04, and a p-value of 0.13, suggesting some variability across studies but not to a concerning degree.

**Figure 2A.**
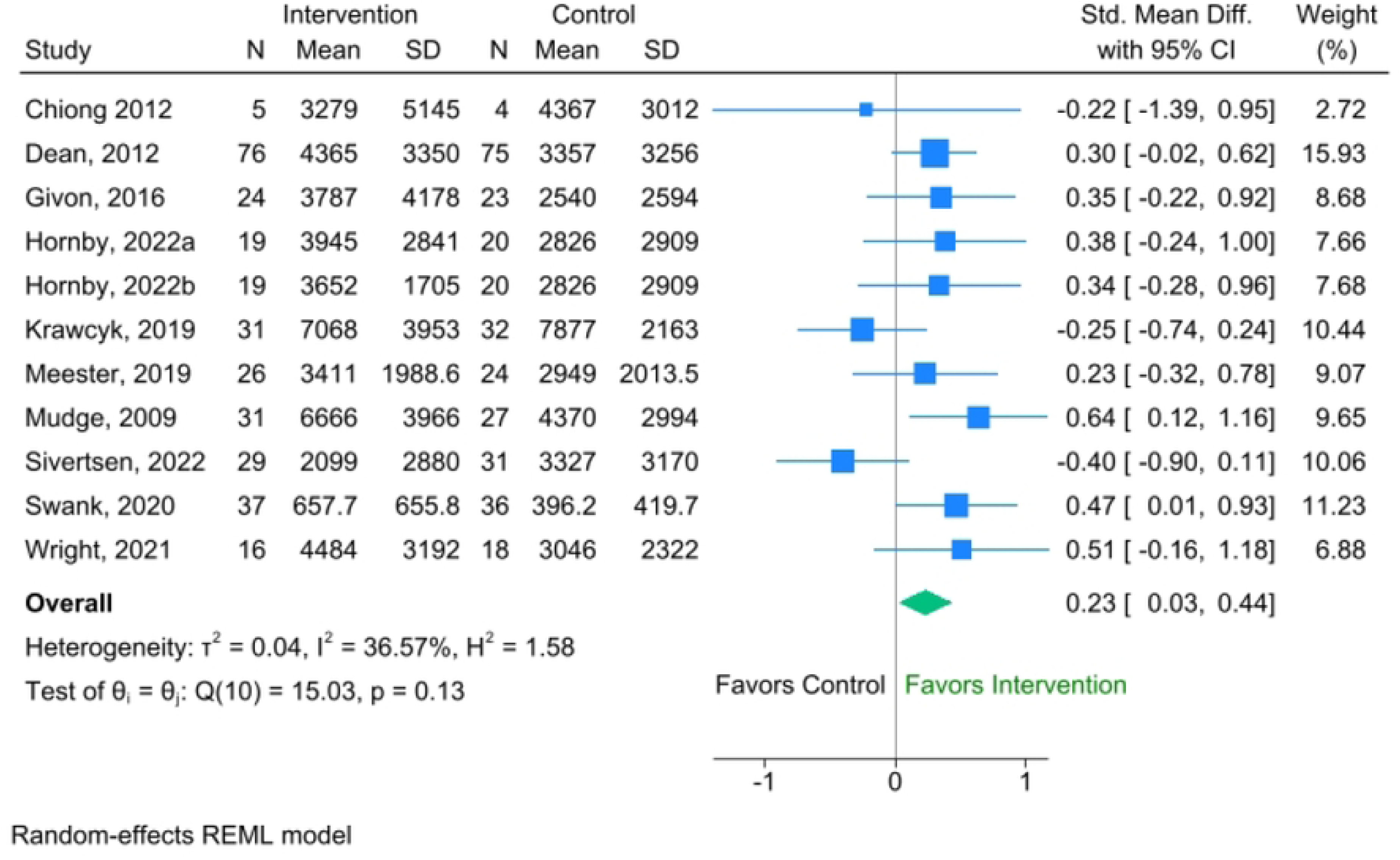
Forest plot for the effectiveness of exercise-only interventions on steps per day

##### Meta-analysis of BCT-only interventions on steps per day

The meta-analysis of BCT-only interventions on daily steps (Figure 2B) included three studies (17,40,41), involving 155 participants in the experimental groups and 162 in the control groups. The pooled analysis using a random effects model revealed an SMD of 0.41 (95% CI: 0.19 to 0.63), indicating a statistically significant positive effect of BCT-only interventions on increasing step count. There was no observed heterogeneity among the included studies, as reflected by an I^2^ value of 0.0%, τ² = 0, and a p-value of 0.76, suggesting consistent findings across all studies.

**Figure 2B.**
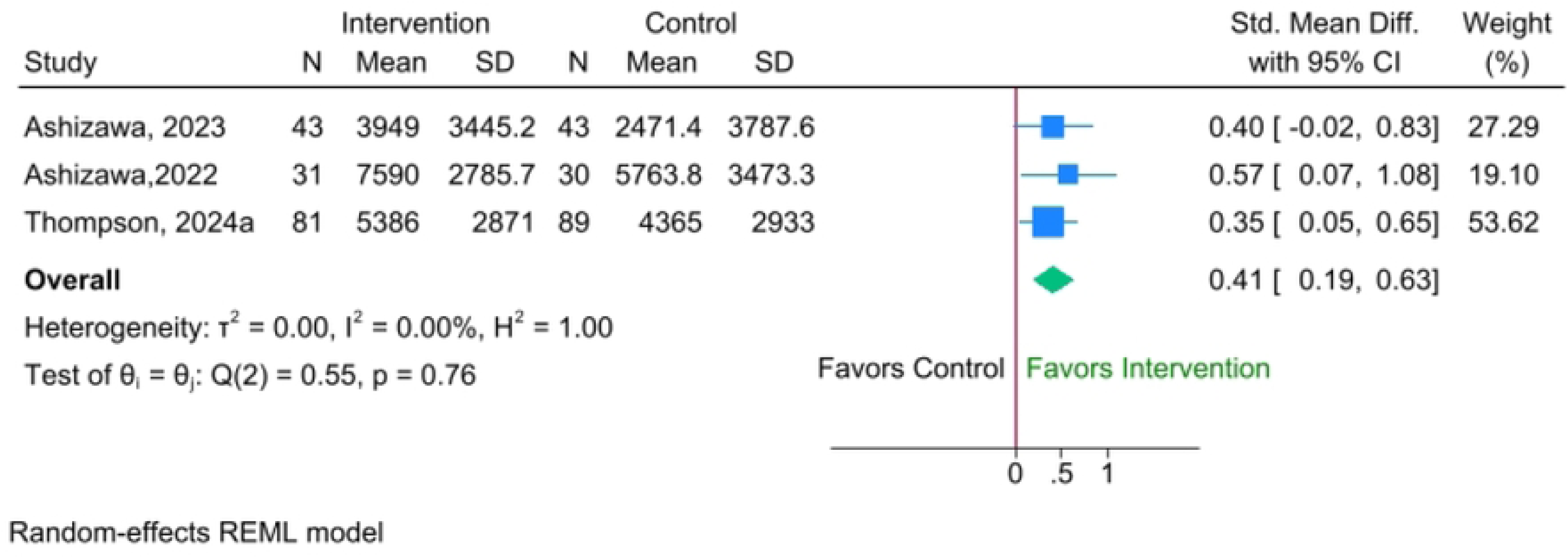
Forest plot for the Effectiveness of BCT-only interventions on steps per day

##### Meta-analysis of combined exercise and BCT interventions on steps

The meta-analysis examining interventions that integrated both exercise and behavior change techniques (BCT) on step count (Figure 2C) encompassed six studies, (17,42,43,47,49,52) involving 234 participants in the intervention groups and 242 in the comparison groups. The analysis, conducted using a random effects model, produced an SMD of 0.39 (95% CI: −0.00 to 0.78). Although the result suggests a potential positive impact, the effect was not statistically significant with very low-quality evidence from studies. Considerable heterogeneity was observed across the studies, indicated by an I^2^ of 76.0%, τ² = 0.18, and a p-value of 0.01, pointing to notable differences in outcomes between studies.

**Figure 2C.**
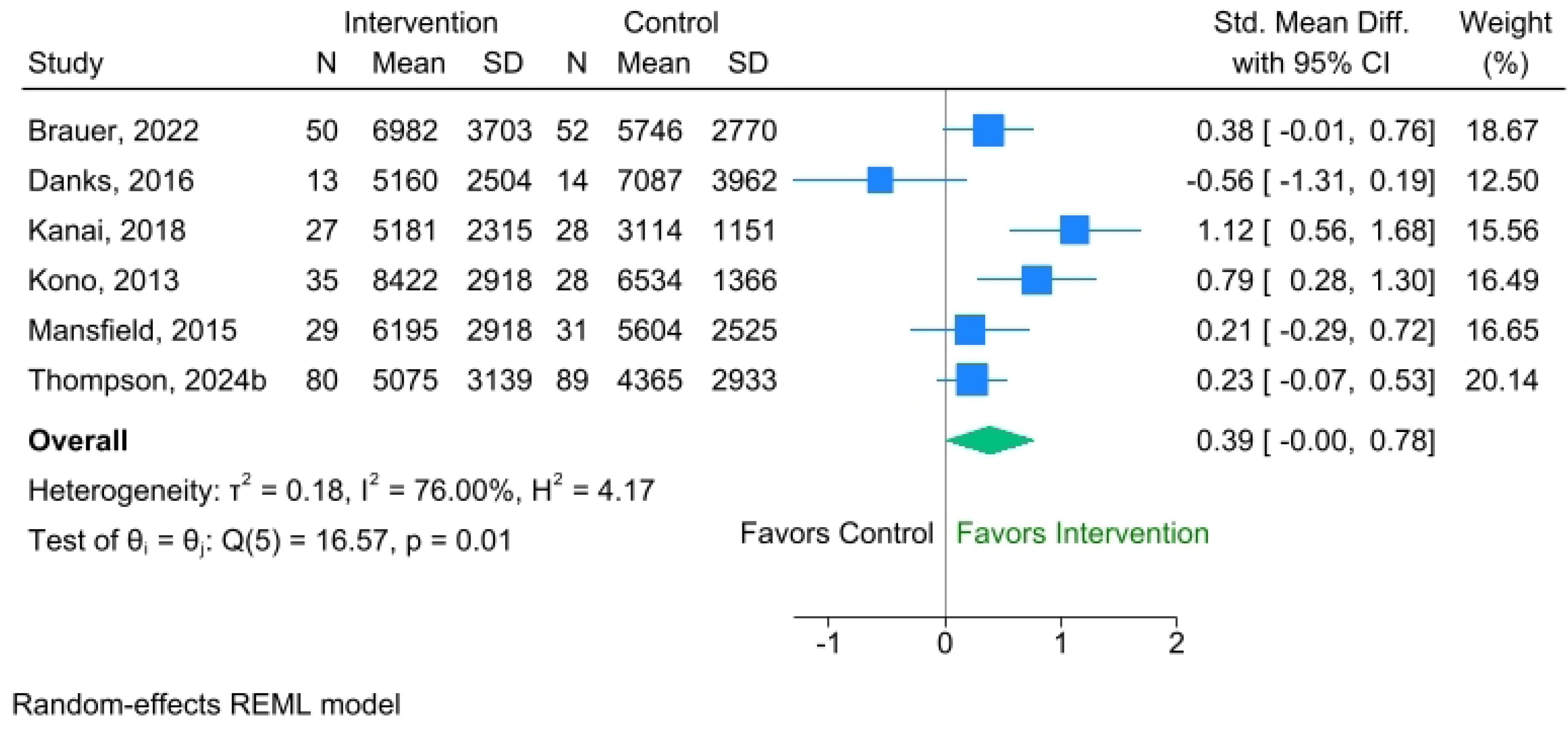
Forest plot for the effectiveness of combined exercise and BCT interventions on steps per day

In summary, exercise-only and BCT-only interventions showed a statistically significant positive effect on steps (SMD 0.23, 0.41, respectively) in the random effects models. The interventions that combined exercise and BCT did not show a significant effect in this model (SMD = 0.39) due to substantial heterogeneity. BCT-only had the largest effect size and no significant heterogeneity.

#### Capacity Outcomes

##### Meta-analysis of the effectiveness of exercise-only interventions on comfortable gait speed

The meta-analysis of exercise-only interventions on the capacity measure of comfortable gait speed included five unique studies (45,48,54,60,62). Because some of these studies had multiple intervention arms, eight comparisons were represented in the forest plot (Figure 3A), comprising 423 participants in the intervention groups and 396 in the control groups. There was moderate-quality evidence of studies and a statistically significant improvement in gait speed among those receiving exercise-only interventions; the pooled SMD was 0.38 (95% CI: 0.19 to 0.57). Moderate heterogeneity was observed across the studies, with an I^2^ value of 35.51%, τ² = 0.02, and a p-value of 0.12, indicating some variability in the results but not to a degree that undermines the overall effect.

**Figure 3A.**
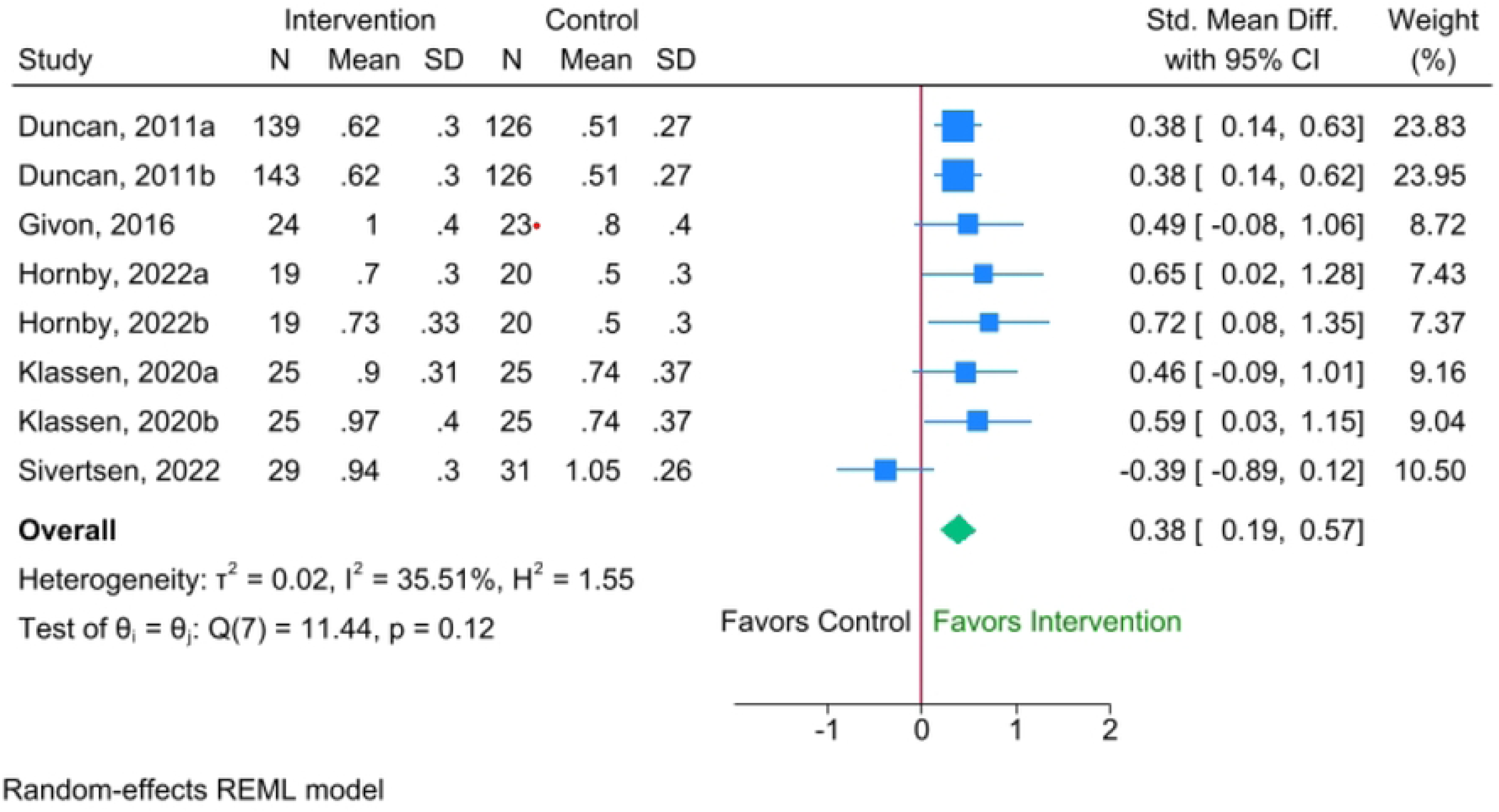
Forest plot for the effectiveness of exercise-only interventions on comfortable gait speed

##### Meta-analysis of the effectiveness of combined exercise and BCT interventions on comfortable gait speed

We did not have sufficient data for BCT-only interventions on comfortable gait speed. The meta-analysis evaluating the effectiveness of combined exercise and BCT on comfortable gait speed (Figure 3B) included four studies, (17,42,43,52) with 188 participants in the intervention groups and 197 in the control groups. The random effects model of very low-quality evidence yielded a standardized mean difference (SMD) of 0.18 (95% CI: −0.16 to 0.52), suggesting a small, positive trend that did not reach statistical significance. Substantial heterogeneity was present among the studies, with an I^2^ value of 60.69%, τ² = 0.053, and a p-value of 0.09, indicating notable variability in the intervention effects across the included trials.

**Figure 3B.**
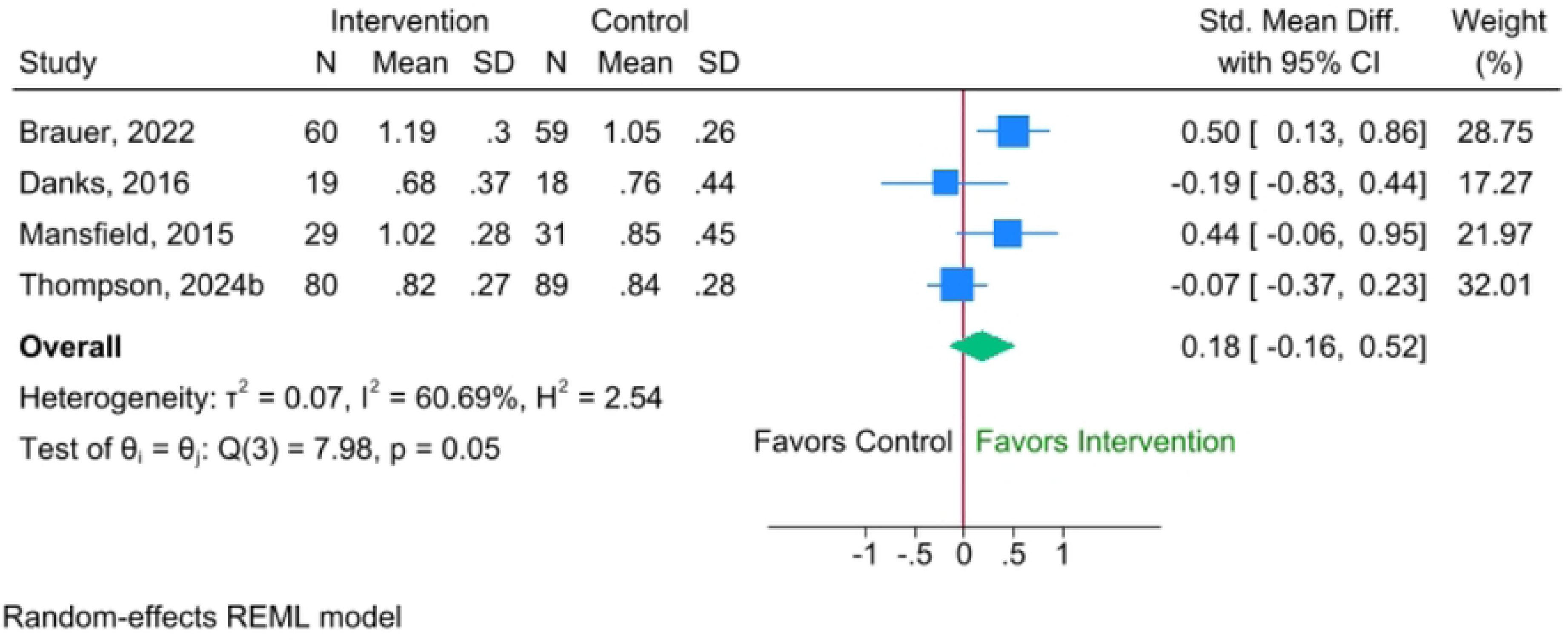
Forest plot for effectiveness of combined exercise and BCT interventions on comfortable gait speed

Overall, for comfortable gait speed, exercise-only interventions demonstrated a statistically significant positive effect (SMD of 0.38) with moderate heterogeneity. In contrast, combined exercise and BCT interventions were not significant (SMD of 0.18) and had substantial heterogeneity, suggesting that the addition of BCTs in these studies did not enhance or consistently contribute to improvements in comfortable gait speed.

##### Meta-analysis of the effectiveness of exercise-only interventions on fastest gait speed

The meta-analysis exploring the effect of exercise-only interventions on fastest gait speed (Figure 4A) included three studies, (54,60,62) comprising 91 participants in the intervention groups and 94 in the control groups. There was very low-quality evidence from studies. The random effects model reported a standardized mean difference (SMD) of 0.43 (95% CI: −0.18 to 1.04), indicating a positive but non-statistically significant effect. Substantial heterogeneity was observed among the studies, with an I^2^ value of 76.8%, τ² = 0.30, and a p-value <0.01, suggesting considerable variation in the outcomes across the included trials.

**Figure 4A.**
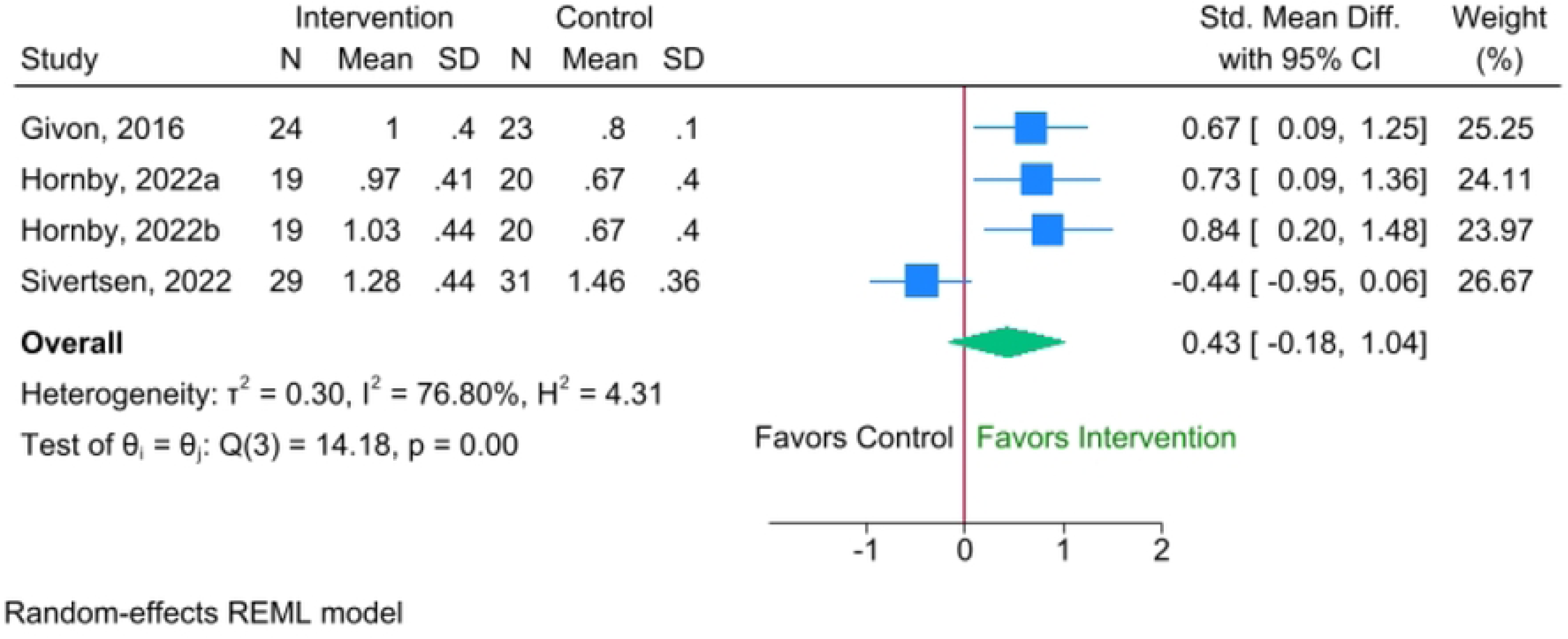
Forest plot for effectiveness of exercise-only interventions on fastest gait speed

##### Meta-analysis of the effectiveness of combined exercise and BCT interventions on fastest gait speed

We did not have sufficient data for BCT-only interventions on fastest gait speed. The meta-analysis examining the effectiveness of combined exercise and BCT interventions on fastest gait speed (Figure 4B) included two studies, (42,43) with 79 participants in the intervention groups and 77 in the control groups. The random effects model produced SMD of 0.28 (95% CI: −0.47 to 1.03), indicating a small, non-statistically significant effect from very low-quality evidence studies. Substantial heterogeneity was detected, with an I^2^ value of 76.57%, τ² = 0.23, and a p-value of 0.04, highlighting considerable variability in the intervention outcomes between studies.

**Figure 4B.**
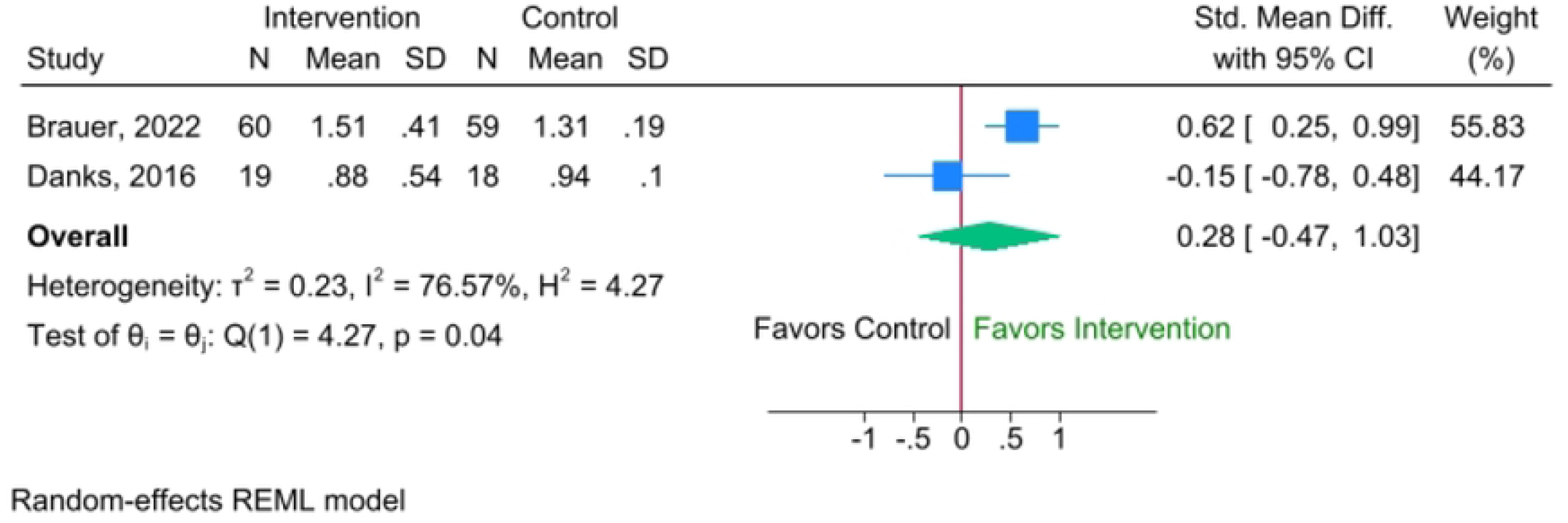
Forest plot for the effectiveness of combined exercise and BCT interventions on fastest gait speed

Overall, for the fastest gait speed, neither exercise-only intervention nor interventions combining exercise and BCT demonstrated a statistically significant effect in the random effects models (SMD of 0.43 and 0.28, respectively). This lack of significance is largely attributable to substantial levels of heterogeneity across studies (I^2^>75% for both).

##### Meta-analysis of the effectiveness of exercise-only interventions on walking endurance

The meta-analysis of exercise-only interventions on walking endurance (Figure 5A) included seven studies, (25,45,48,50,53,60,65) involving a total of 487 participants in the experimental groups and 453 in the control groups. The random effects model showed a standardized mean difference (SMD) of 0.39 (95% CI: 0.26 to 0.52), indicating a statistically significant improvement in walking endurance from moderate quality evidence studies. Heterogeneity among the studies was low, with an I^2^ of 0%, τ² of 0.00, and a p-value of 0.42, suggesting consistent results across the included trials.

**Figure 5A.**
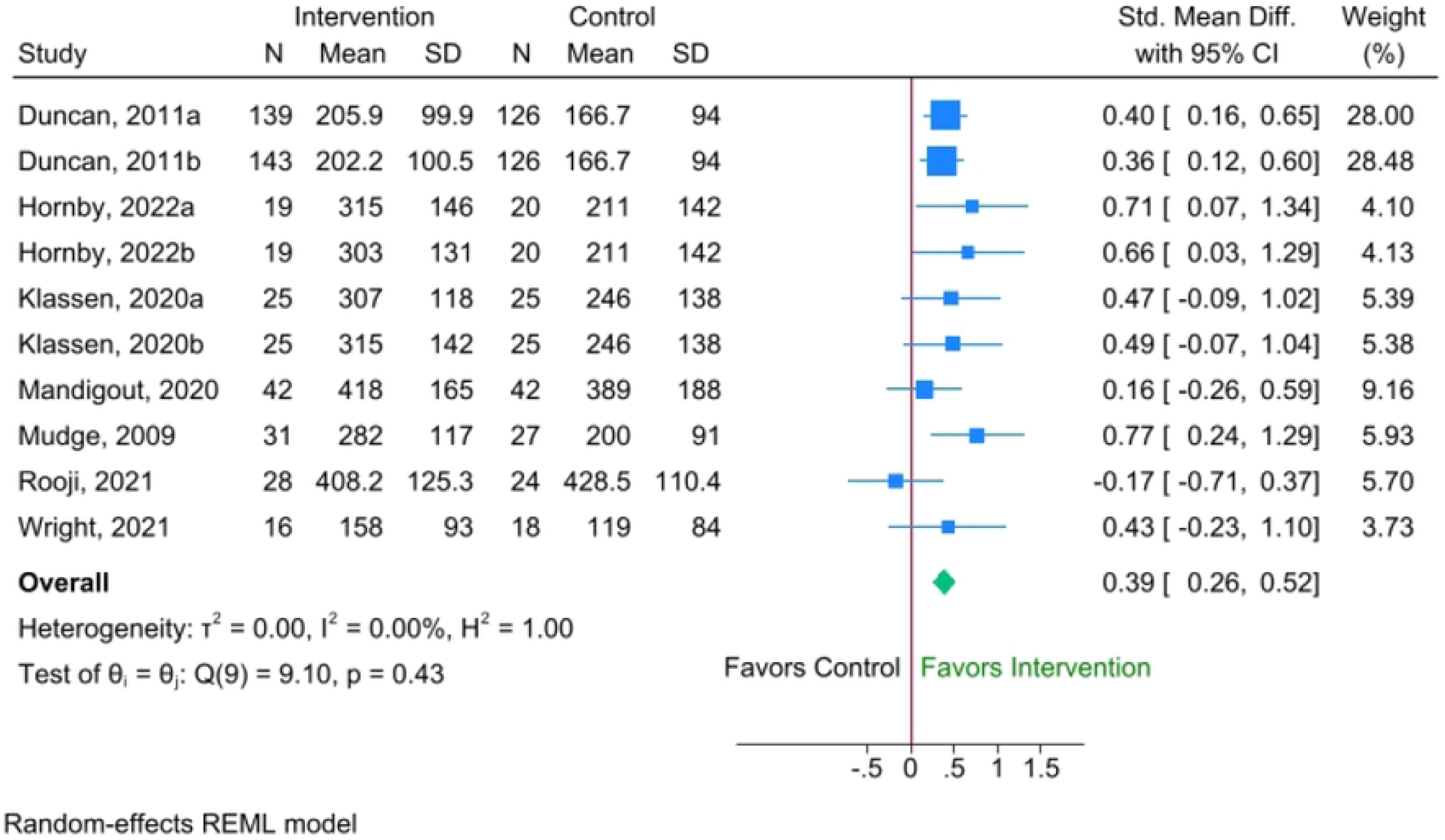
Forest plot for the effectiveness of exercise-only interventions on walking endurance

##### Meta-analysis of BCT-only interventions on walking endurance

The meta-analysis evaluating the effectiveness of BCT-only interventions on walking endurance (Figure 5B) included two studies, (17,55) comprising 123 participants in the intervention groups and 130 in the control groups. There were very low-quality evidence studies, and the random effects model yielded an SMD of −0.03 (95% CI: −0.56 to 0.50), indicating no significant effect on walking endurance. Substantial heterogeneity was observed between the studies, with an I^2^ value of 75.78%, τ² = 0.11, and a p-value of 0.04, reflecting considerable variation in outcomes.

**Figure 5B.**
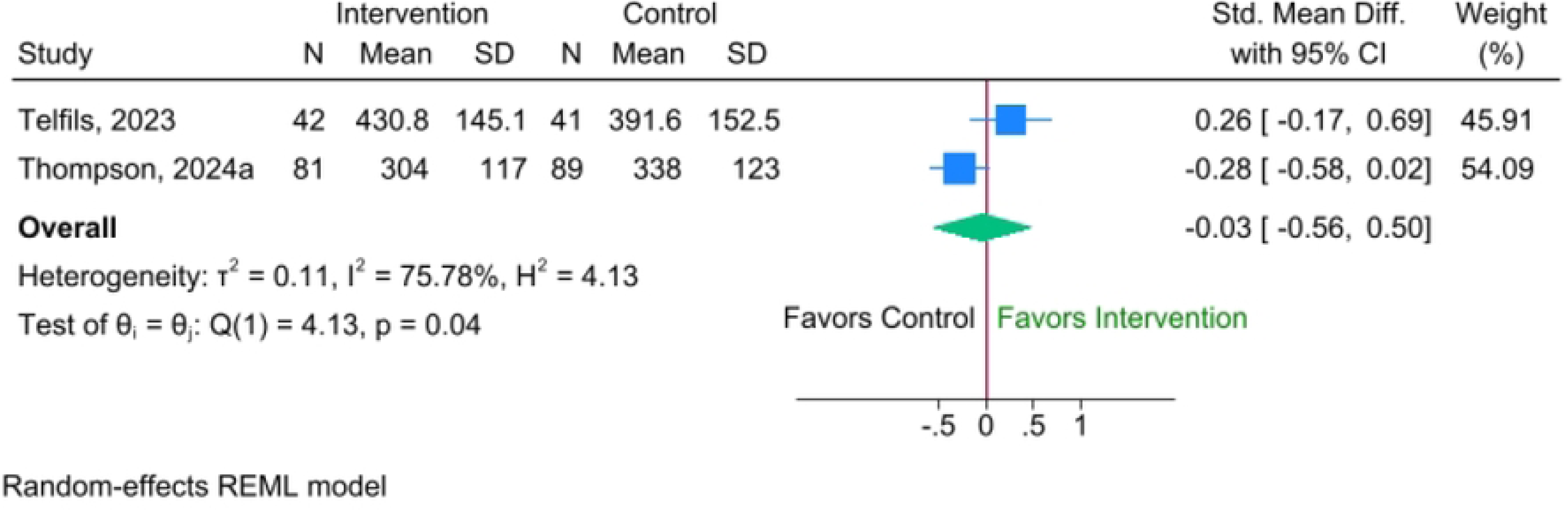
Forest plot for the effectiveness of BCT-only interventions on walking endurance

##### Meta-analysis of combined exercise and BCT interventions on walking endurance

The meta-analysis assessing the effectiveness of combined exercise and BCT interventions on walking endurance (Figure 5C) included two studies (17,43) involving 93 participants in the intervention groups and 103 in the control groups. There were low-quality evidence studies, and the random effects model produced a standardized mean difference (SMD) of −0.10 (95% CI: −0.38 to 0.18), indicating a small, non-statistically significant effect. This suggests that, based on current evidence, combined interventions may not lead to meaningful improvements in walking endurance. There was no heterogeneity, with an I^2^ of 0.0%, τ^2^ of 0.0, and a p-value of 0.77.

**Figure 5C.**
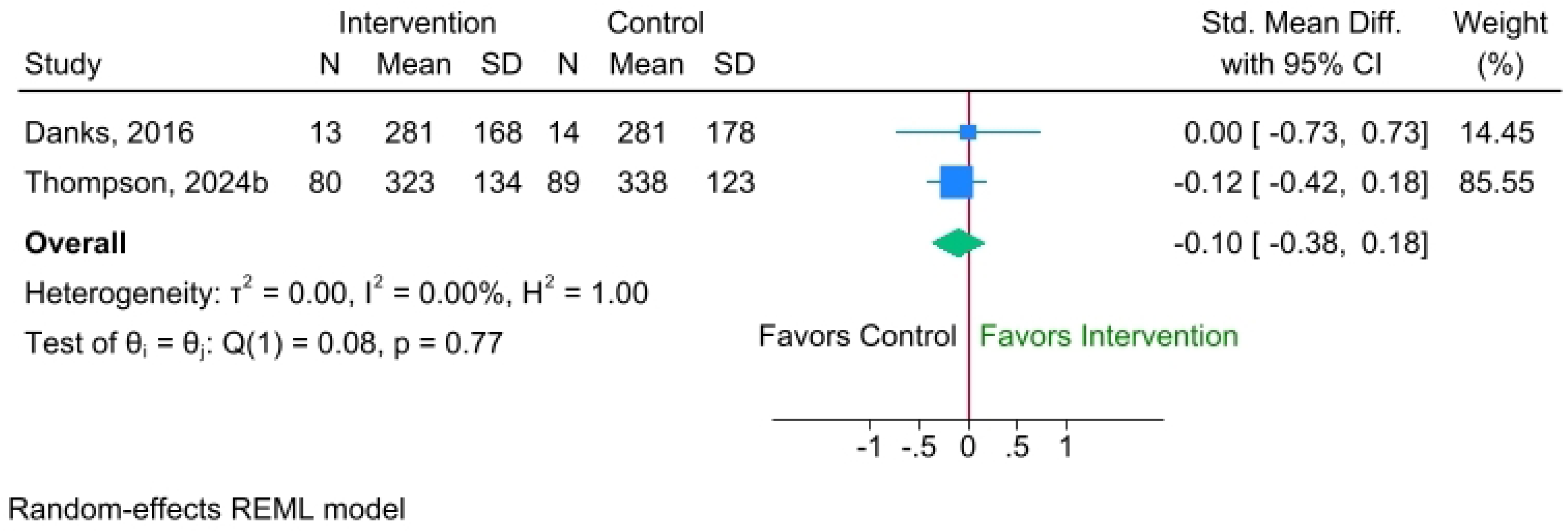
Forest plot for effectiveness of combined exercise and BCT interventions on walking endurance

Overall, for walking endurance, exercise-only interventions demonstrated a statistically significant positive effect SMD of 0.39 (95% CI: 0.26 to 0.52) in the random effects models, with no heterogeneity (I^2^ of 0%, τ^2^ of 0.00, and a p = 0.42). In contrast, neither BCT-only nor combined interventions showed a statistically significant effect. The BCT-only intervention exhibited substantial heterogeneity, while the combined intervention showed no heterogeneity but did not reach statistical significance. This suggests that for improving endurance, exercise alone is the most effective intervention among the three categories. Adding BCTs, either as a standalone or combined approach, did not yield statistically significant improvements in 6MWT in the analyzed studies.

## 4. Discussion

Current rehabilitation paradigms for individuals’ post-stroke often prioritize capacity-based outcomes, such as gait speed and the 6MWT, over real-world performance measures like daily step counts. Yet, growing evidence suggests that improvements in capacity do not always translate into increased daily PA (3,17,24,25). This disconnect between what individuals *can* do in clinical settings and what they *actually* do in daily life represents a significant gap in post-stroke rehabilitation. This systematic review and meta-analysis addressed this issue by evaluating the effects of physical rehabilitation interventions - exercise-only, BCT-only, and combined Exercise + BCT-on daily activity behaviours post-stroke. Our findings and quality of evidence showed that exercise-only interventions significantly improved both walking endurance and comfortable gait speed, consistent with prior work demonstrating that aerobic and task-specific training enhance cardiovascular fitness and functional mobility post-stroke (43,60,66–70). However, these interventions produced an increase in daily steps, contrasting with a previous meta-analysis by Oliveira et al. (28), which found no significant effects of exercise alone on step count. Notably, Oliveira et al. (28) did not include capacity outcomes in their synthesis and focused only on performance, whereas our review included both capacity and performance outcomes, enabling a more comprehensive assessment. Furthermore, the current meta-analysis included studies across different recovery stages compared to that analysis, which was limited to acute/subacute stroke studies (28).

To our knowledge, no previous systematic review has directly compared the effects of physical rehabilitation interventions on both performance and capacity outcomes following stroke. We found that BCT-only interventions had a higher magnitude of effect on daily steps (SMD = 0.41; 95% CI: 0.03 to 0.44) compared to exercise-only interventions, with no heterogeneity across studies. This supports the theoretical premise that behavioural interventions such as goal setting, feedback, self-monitoring, and action planning are effective in promoting real-world increases in performance (71–73). These findings are consistent with prior research demonstrating the effectiveness of behavioral strategies in enhancing activity levels across various chronic disease populations, including stroke survivors (15,17,32,74,75). Given the high prevalence of physical inactivity after stroke and its strong association with adverse outcomes such as recurrent stroke, disability, and mortality (26,76,77), our results underscore the critical role of integrating BCTs into rehabilitation frameworks. Moreover, current evidence suggests that even modest increases in PA—such as an additional 1,000 steps per day—can reduce all-cause mortality by 15% to 23% (78,79), highlighting the significant public health implications of these interventions.

Interestingly, while several individual studies of combined Exercise + BCT interventions showed positive effects on daily steps (17,42,47,49,52), our pooled analysis which included 6 studies did not result in a statistically significant overall benefit (SMD = 0.39; 95% CI: −0.00 to 0.78) and revealed substantial heterogeneity (I^2^ = 76%). A key factor contributing to this result appears to be the inclusion of the study by Danks et al. (43) which reported a negative effect size (SMD = −0.56; 95% CI: −1.31 to 0.19) that likely reduced the overall pooled estimate. In contrast, Oliveira et al. (28) found a significant effect of combined Exercise and BCT interventions on daily steps in individuals with acute and subacute stroke (SMD = 0.65; 95% CI: 0.23 to 1.06; p = .002), with considerable heterogeneity (χ² = 8.32; p = .04; I^2^ = 64%). Notably, their analysis did not include the study by Danks et al. (43) which may partly explain the more favourable pooled estimate. These contrasting findings underscore how methodological decisions, including study selection, timing of intervention (acute vs. chronic stroke), and intervention characteristics, can influence meta-analytic results. Moreover, combining exercise and behavioural interventions may introduce complexity that undermines their individual effectiveness especially if the interventions are not specifically tailored to the specific outcome of interest.

Regarding gait speed, our meta-analysis found that exercise-only interventions significantly improved comfortable gait speed (SMD = 0.38), in line with findings from previous trials (17,60,62) and reinforcing the value of task-specific training. In contrast, interventions combining BCT and exercise showed no significant improvement and considerable heterogeneity. Fastest gait speed was not significant in any intervention category—likely due to wide variability in protocols, populations, and measurement approaches.

For walking endurance (measured by the 6MWT), exercise-only interventions again showed a statistically significant benefit (SMD = 0.39), with no heterogeneity. In contrast, BCT-only and combined interventions did not produce significant changes. This pattern is logical, as endurance improvements are closely tied to physiological training thresholds that are unlikely to be influenced by BCTs alone. These results align with the motor learning and overload principles underpinning physical rehabilitation: without sufficient practice intensity or cardiovascular challenge, physiological adaptations are unlikely to occur (19). The lack of benefit from BCT-only interventions for capacity outcomes highlights the importance of aligning intervention targets with outcome domains.

A particularly noteworthy finding was the underperformance of combined Exercise + BCT interventions across most outcomes. While intuitively appealing, these interventions may have suffered from implementation challenges, mismatched content, or intervention overload. Some BCTs might overlap with strategies already embedded in structured exercise programs, offering no additional benefit. Furthermore, without detailed reporting of intervention fidelity and participant engagement, it’s difficult to determine whether the intended synergies were achieved. High heterogeneity in this group further suggests a lack of standardization, making it difficult to draw strong conclusions.

In summary, this review highlights that targeted intervention strategies may be more effective than combined approaches when aiming to improve specific outcomes post-stroke. Exercise-only interventions appear best suited for enhancing capacity outcomes such as gait speed and endurance, while BCT-only interventions are particularly effective for increasing real-world performance, as measured by daily steps. These findings have important clinical implications, suggesting that rehabilitation programs should consider tailoring interventions based on the specific outcomes of interest rather than assuming that more complex or combined approaches are inherently superior.

### Clinical Implications and Future Directions

For clinicians and rehabilitation teams, these results emphasize the need to tailor interventions based on the desired outcome. If the goal is to enhance real-world PA, BCT-only interventions such as goal setting, self-monitoring, and feedback may be particularly effective and practical to implement, especially in community settings. Conversely, to improve capacity-based outcomes like gait speed and endurance, structured exercise-based interventions remain essential. The underperformance of combined interventions suggests that simply layering behavioural strategies onto exercise may not yield added benefit unless both components are delivered with high fidelity and strategic alignment. Clinicians should also be aware that increasing capacity does not automatically lead to improved performance, underscoring the need to assess and target both domains separately.

### Limitations

This review has several limitations. First, although the search was comprehensive, some relevant studies may have been missed due to variations in terminology or limited reporting of intervention content. Second, there was considerable heterogeneity in study design, participant characteristics (e.g., stroke chronicity), intervention intensity, and outcome measurement, particularly within the combined intervention group. Third, the classification of BCTs relied on reported descriptions, which may not always reflect actual fidelity or quality of delivery. Additionally, publication bias cannot be ruled out, and the overall number of studies in some subgroups (e.g., BCT-only for gait speed) was limited, reducing the power to detect effects in those domains. Finally, only quantitative outcomes were included; qualitative insights on acceptability, feasibility, or patient experience were beyond the scope of this review.

## 5. Conclusions

This systematic review and meta-analysis provide the first comprehensive comparison of the effects of exercise-only, BCT-only, and combined Exercise + BCT interventions on both performance (daily steps) and capacity (gait speed and endurance) outcomes in individuals’ post-stroke. Our findings reveal that while exercise-only interventions consistently improve capacity outcomes such as gait speed and endurance, BCT-only interventions yielded the most reliable improvements in daily step count, a critical performance measure. Surprisingly, combined interventions did not consistently outperform single-component strategies and were associated with substantial heterogeneity. These findings highlight the importance of matching intervention strategies to specific rehabilitation goals and suggest that “more” is not always “better” when it comes to multimodal rehabilitation design.

## Data Availability

All relevant data are within the manuscript and its Supporting Information files.

## 7. Supporting Information

S1 Table. This is the S1 Table PRISMA Checklist

S1 File. This is the S1 File Search Strategy

S2 File: This is the S2 File Data Extraction Sheet

S2 Table: This is the S2 Table GRADE Certainty of Evidence

